# Apathy and Physical Activity: A Systematic Review and Meta-Analysis

**DOI:** 10.1101/2024.05.01.24306712

**Authors:** Ata Farajzadeh, Alexe Hébert, Ian M. Lahart, Martin Bilodeau, Matthieu P. Boisgontier

## Abstract

**Purpose:** Patient motivation is essential for the success of their rehabilitation. Apathy, a state of primary motivational deficiency, may therefore hinder physiotherapists’ interventions, such as those aimed at increasing patients’ physical activity. This study aims to examine the relationship between apathy and physical activity and to identify the factors that influence this relationship.

**Methods:** Six databases were searched for articles including both a measure of apathy and physical activity. Two reviewers screened articles for inclusion, assessed risk of bias, and extracted data from each study. Pearson product-moment correlations were pooled from eligible studies using the generic inverse pooling and random effects method to examine the relationship between apathy and physical activity. Subgroup meta-analyses were conducted to examine the differences between studies that included participants with different health conditions and used different types of physical activity outcomes and apathy measures. Meta-regressions were conducted to examine the moderating effects of age and gender.

**Results:** Twenty-eight articles were included in the systematic review and 22 studies (n = 12,541 participants) in the main meta-analysis. Results showed a negative correlation between apathy and physical activity (r = -0.13; 95% CI: -0.18 to -0.09; p < 0.0001 based on Pearson’s r values; r = - 0.40; 95% CI: -0.68 to -0.02; p = 0.043 based on Spearman’s rho values). A subgroup meta-analysis revealed that the correlation was statistically significant in patients with Parkinson’s disease and in older adults who were healthy, depressed, fallers, or had mild cognitive impairment. A meta-regression showed an effect of age, with a stronger correlation between apathy and physical activity in older adults compared to younger adults (p = 0.003).

**Conclusion:** Our results suggest that higher levels of apathy are associated with lower levels of physical activity and that this negative association is stronger with age. Therefore, apathy may limit exercise therapy efficacy and carry prognostic implications for patients whose condition requires physical activity.

## 1. Introduction

Psychological factors are recognized as essential to quality rehabilitation interventions (1). Motivation, for example, defined as the hypothetical construct used to describe the internal and external forces that produce the initiation, direction, intensity, and persistence of behavior (2), is a parameter that physiotherapists should consider in their intervention, as it influences the patient’s ability to adhere to the prescribed rehabilitation program (3). While motivation can vary from patient to patient within a physiological range, it can also be pathologically altered and lead to ineffective care (4). An example of such alteration is apathy, which is conceptualized as a behavioral disorder (5,6).

In the early 1990s, apathy began to be considered a syndrome rather than a mere symptom of another condition, and was defined as a state of primary motivational impairment characterized by reduced goal-directed behaviors (e.g., lack of productivity, effort, initiative, and perseverance), reduced goal-directed cognitions (e.g., lack of interest in learning new things, in new experiences, lack of concern about one’s personal and health problems, reduced socialization), and reduced emotional concomitances of behavior (e.g., flat affect, emotional indifference) (5,6). To date, the dominant theoretical frameworks still consider three subtypes of apathy: behavioral, cognitive, and emotional (7,8). Levy and Dubois (9) proposed that behavioral apathy is related to difficulties in elaborating the plan of actions necessary for behavior. Cognitive apathy is the inability to self-activate thoughts or self-initiate actions. Emotional apathy is the inability to establish the linkage between emotional signals and behavior (9). A fourth subtype, social apathy, has recently been proposed but requires further research (10).

Apathy occurs across a wide range of neurological and psychiatric disorders (11), including dementia (54%) (12), schizophrenia (47-53%) (13–15), Parkinson’s disease (40%) (16), and stroke (33%) (17). Apathy has also been observed in healthy young adults (1.5%) (18) and older adults with normal cognitive function (2-7%) (19–23). Apathy is associated with lower cognitive performance in older adults with normal cognitive functioning (20), cognitive decline (mild cognitive impairment) (21), transition to dementia (24,25), and dementia severity (26,27). Therefore, apathy could be considered a marker of impending cognitive decline and future risk for dementia (20). Moreover, apathy has been associated with frailty (26), functional decline (27–30), poorer quality of life (18,23,33), higher mortality (34,35), and higher healthcare costs (36).

In addition, some studies suggest that apathy can negatively affect physical activity (27–40), which includes exercise, sports, active travel (cycling, walking), household chores, and work-related physical activity (41). This potential effect of apathy is worth investigating since physical activity is now widely recognized as one of the top contributors to physical and mental health, improving cognitive functioning (42), and reducing rates of cardiovascular disease (43), cancer (44), hypertension (45), diabetes (46), obesity (47), depression (48), and functional dependence (48–50). Despite these benefits, one in four adults fails to meet the recommended levels of physical activity (41). While the motivational impairment that defines apathy is likely to reduce the engagement in physical activity, there are some discrepancies in the literature, with multiple articles showing no evidence of such relationship (51,52) or even supporting the opposite association (53).

The main objective of this study was to conduct a systematic review and meta-analysis of the direct relationship between apathy and physical activity. We hypothesized that levels of apathy would be negatively associated with levels of physical activity. Since studies assessing this relationship used different methods and outcome measures in a variety of health conditions, we explored whether apathy measure, physical activity measure (e.g., accelerometers, pedometers, questionnaires), physical activity outcome (e.g., total physical activity, moderate or vigorous physical activity, steps per day), and health status influenced the results. Additionally, since older age and being a woman have been associated with reduced physical activity (54) and higher apathy (55,56), we explored whether these demographics influenced the results.

## 2. Methods

### 2.1. Search Strategy

This review is reported in accordance with the Preferred Reporting Items for Systematic Reviews and Meta-Analyses (PRISMA) guidelines (57). Potential studies were identified by searching the MEDLINE (via PubMed), PsycINFO, Web of Science, Embase, SPORTDiscus, and CINAHL databases. The search terms used to identify relevant studies were variants of physical activity (e.g., physical activity, physical education, training, physical fitness, exercise, sport, walk) and apathy (e.g., abulia, apathetic, amotivation, avolition, neuropsychiatric inventory, NPI (NeuroPsychiatry Inventory), emotional indifference, frontal lobe personality scale, Lille apathy rating scale, LARS (Lille apathy rating scale), dementia apathy interview and rating, DAIR (Dementia Apathy Interview and Rating), frontal system behavior scale, FrSBe (Frontal System Behavior scale), key behaviors change inventory, KBCI (Key Behaviors Change Inventory), apathy evaluation scale, apathy scale, and irritability apathy scale (Supplemental Material 1). Articles were searched up to October 10, 2023, with no limitation on the start date. A filter was used to limit the search to studies published in English. To reduce literature bias, this systematic review was pre-registered in PROSPERO (CRD42023492162) (58).

### 2.2. Eligibility Criteria and Study Selection

#### 2.2.1. Inclusion Criteria

To be included in this systematic review, articles had to 1) be published in a peer-reviewed journal, 2) be written in English, 3) report original data collected from human participants, 4) include at least one self-reported measure of apathy and at least one measure of physical activity, and 5) formally test the association between these two variables (e.g., reporting Pearson’s r or Spearman’s rho correlation coefficient). The physical activity measure could be a self-reported measure or device-based measure (e.g., accelerometry) of the level of physical activity. Cohort studies, baseline data from clinical trials, and cross-sectional studies were included in this review. The inclusion criteria did not impose any specific age or gender restrictions.

#### 2.2.2. Exclusion Criteria

Studies were excluded if they were 1) published as a book chapter, study protocol, or conference abstract, or 2) based on laboratory-based measures of physical fitness (e.g., maximal muscle force, VĳO_2_ max) and not on a measure of physical activity.

#### 2.2.3. Study Selection

Articles were screened using the Covidence systematic review software (Veritas Health Innovation, Melbourne, Australia), a web-based collaborative software platform that streamlines the production of systematic reviews. After duplicates were removed, titles and abstracts were independently reviewed by two reviewers who authored this article (AF, AH) according to the inclusion and exclusion criteria. If there was any doubt about the inclusion or exclusion of an article, the full text was reviewed by the two reviewers. In addition, reference screening and forward citation tracking were performed on the included articles. Disagreements between the two reviewers were resolved by consensus among four reviewers (AF, AH, MB, MPB).

### 2.3. Data Extraction

Data extracted from the selected articles included first author’s name, article title, publication year, number of participants, number of men and women, mean age and range, health status, type of apathy measure, level of apathy, type of physical activity measure and outcome, level of physical activity, as well as statistical estimates, and significance of the association between apathy and physical activity. The data extraction form was developed by the authors and was previously tested in a systematic review (59,60).

### 2.4. Bias Assessment

The included studies were assessed for methodological quality using 10 questions from the Quality Assessment Tool for Observational Cohort and Cross-Sectional Studies (61). Two reviewers (AF, AH) independently answered these questions for each study, with a third reviewer (MPB) involved in case of disagreement.

### 2.5. Meta-Analysis

All analyses were conducted in RStudio integrated development environment (IDE) (2023.06.1+524, ‘Mountain Hydrangea’ release) for R software environment (62) using the ‘meta’ (63), ‘metasens’ (64), and ‘metafor’ (65,66) R packages.

#### 2.5.1. Main Meta-Analysis

In the main meta-analysis, we pooled Pearson product-moment correlations from eligible studies to examine the relationship between apathy and physical activity. When a correlation coefficient was not reported, but the exact p value (or t value) and sample size were available, and it was possible to know the sign of the correlation, the Pearson’s r value was computed using an ad-hoc R code (Supplemental R Script 1) using the ‘qt’ function of the R ‘MASS’ package to find the critical value of t (67,68). For the studies that reported a relative p-value < 0.001 instead of an exact p-value, we used a p-value of 0.0009 to estimate an approximate r value. When exact p-values were not reported in an article, but the sample size (n) and Pearson’s correlation coefficient were available, the exact p-value was computed using an ad-hoc R code (Supplemental R Script 2). Correlations were pooled using the generic inverse pooling method via the ‘metacor’ function in the R ‘meta’ package (63). This function automatically performs a necessary Fisher’s z-transformation on the original, untransformed correlations prior to pooling. The ‘metacor’ function reconverts the pooled association back to its original form for ease of interpretation. We anticipated considerable between-study heterogeneity and therefore used a random-effects model to pool correlations. The restricted maximum likelihood (RML) estimator (69) was used to calculate the heterogeneity variance Tau^2^. In addition to Tau^2^, to quantify between-study heterogeneity, we report the I^2^ statistic, which provides the percentage of variability in the correlations that is not caused by sampling error (70). The I^2^ statistic was interpreted as: 0-40%, may not be important; 30-60%, may represent moderate heterogeneity; 50-90%, may represent substantial heterogeneity; and 75-100%, may represent considerable heterogeneity. To reduce the risk of false positives, we used a Knapp-Hartung adjustment (71) to calculate the confidence interval around the pooled association. We also report the prediction interval, which provides a range within which we can expect the associations of future studies to fall based on the current evidence. If significant, the pooled correlation was interpreted using Cohen’s conventions (72): r ≈ -0.10, small negative correlation; r ≈ -0.30, moderate negative correlation; r ≈ -0.50, large negative correlation.

#### 2.5.2. Publication Bias Assessment

Publication bias refers to the fact that studies with positive or significant results are more likely to be published, while studies with negative or non-significant results are less likely to appear in the literature. Therefore, publication bias can affect the validity and generalization of conclusions of a meta-analysis (73). Here, publication bias was assessed using a funnel plot, which is a scatter plot of the studies’ effect size expressed as the Fisher’s z transformed correlation on the x-axis against a measure of their standard error (which is indicative of precision of the study’s effect size) on the y-axis. Egger’s regression was used to formally test funnel plot’s asymmetry (74). Because the probability of obtaining significant results rises with a larger sample size, publication bias will disproportionately affect small studies. Rücker’s limit meta-analysis method explicitly accounts for the publication bias due to small studies (75) and was used to compute a bias-corrected estimate of the true effect size. More specifically, the model underlying Rücker’s method is an extended random-effects model that takes account of possible small study effects by allowing the tested effect to systematically depend on the standard error (Supplemental Material 2).

Another method used to assess publication bias was p-curve analysis (76). When the null hypothesis is true (i.e., there is no true effect), p-values are assumed to follow a uniform distribution: Highly significant effects (e.g., p = 0.01) are as likely as barely significant effects (e.g., p = 0.049). However, when the null hypothesis is false (i.e., there is a true effect in our data), p-values are assumed to follow a right-skewed distribution: Highly significant effects are more likely than barely significant effects. A left-skewed distribution would suggest that some studies used statistical tests to find significant results in ways that may not be reproducible or generalizable (i.e., p-hacking).

#### 2.5.3. Secondary Meta-Analysis

A secondary meta-analysis was conducted using the same approach as in section 2.5.1, but based on Spearman’s rho values, to further test the relationship between apathy and physical activity. We chose not to compare r and rho values, or any transformed versions of them, in the same meta-analysis because they do not reflect the same property of the data. In the secondary meta-analysis, some studies reported multiple outcomes for the same sample of participants. Therefore, correlation estimates were nested within studies using the ‘cluster’ argument to account for the dependencies between these estimates, resulting in a three-level meta-analysis (level 1: participants, level 2: correlation estimates, level 3: studies). The distribution of variance across levels was assessed using the multilevel version of I^2^ (77). The performance of the 2-level and 3-level meta-analyses was assessed and compared using the ‘metafor’ R package (65,66).

#### 2.5.4. Subgroup Meta-Analyses and Meta-Regressions

Subgroup meta-analyses were conducted to examine the differences in correlations between studies that included participants with different health conditions, used different types of physical activity outcomes, and used different types of apathy measures.

Meta-regressions were conducted to examine if the average age of participants or the proportion of women in a study predicted the reported correlation between apathy and physical activity. Another meta-regression was used as a sensitivity analysis to examine whether the quality of the studies affected the correlation.

## 3. Results

### 3.1. Literature Search

The primary search identified 6,950 potentially relevant articles from the six databases (Figure 1), including 2,025 duplicates. Of the 4,925 articles screened, 3,441 were unrelated to apathy or physical activity. Of the remaining 1,484 articles, 65 met the criteria for further investigation and potential inclusion. To increase the completeness of our review, we emailed the corresponding authors of 42 articles that did not formally test the association between the apathy and physical activity to request the correlation estimate or raw data so we could compute this estimate ourselves. Six authors replied: one provided raw data (78), one provided the Pearson’s correlation coefficient (79), and four replied that they would contact their co-authors but did not get back to us (80–83). Reference screening and forward citation tracking identified three additional studies (84–86), of which one provided the Pearson correlation upon request (84). Detailed reasons for exclusion at each stage are provided in the PRISMA flow diagram (Figure 1). Ultimately, 28 articles were included in the systematic review. Two of these articles, published by the same group, had similar methods and partly overlapping recruitment periods, suggesting that some participants may have been included in both studies (52,87). Quality score of studies ranged from 6 to 10 out of 10, with a mean ± standard deviation of 8.4 ± 1.2 (Table 1).

**Figure 1.**
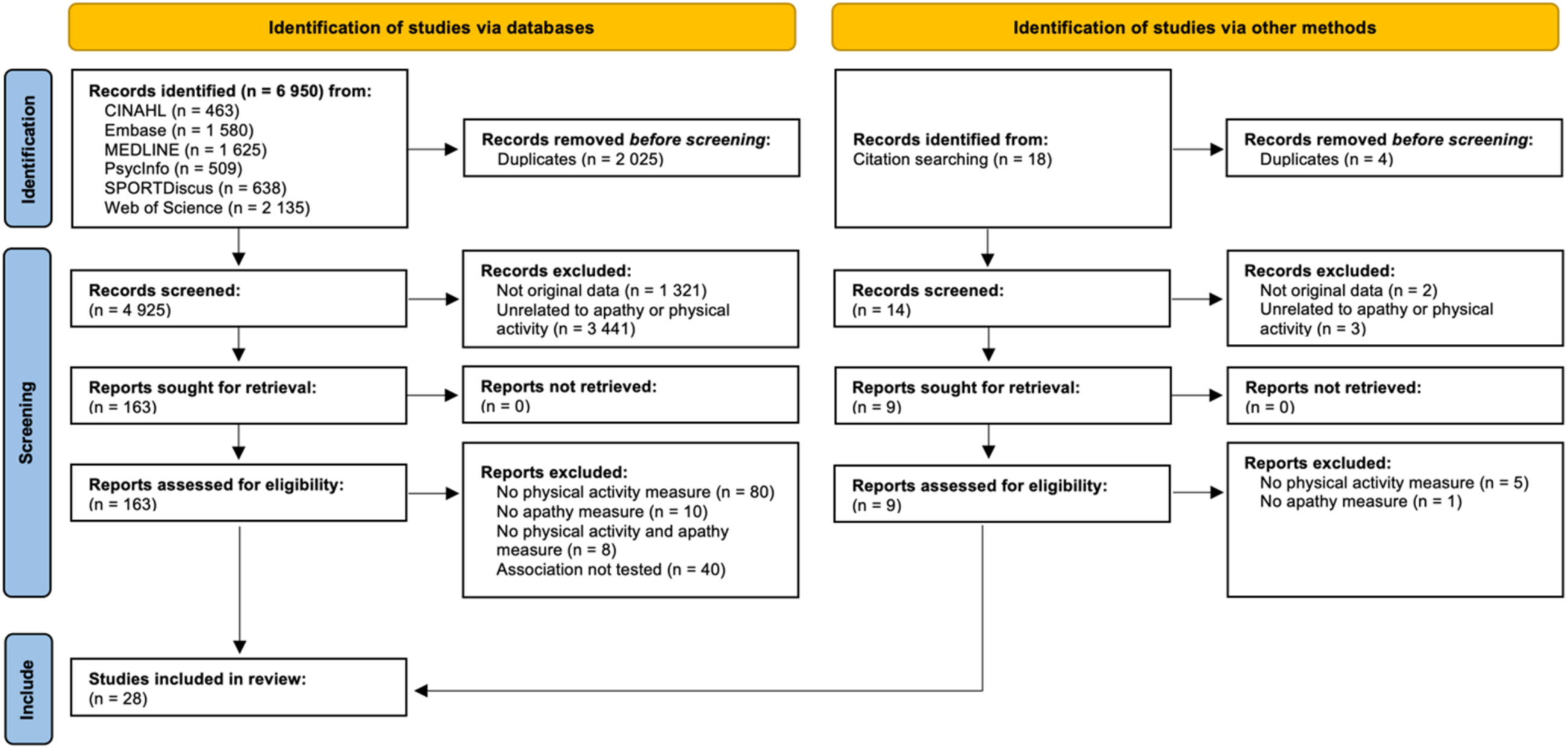
PRISMA 2020 flow diagram.

**Table 1.**
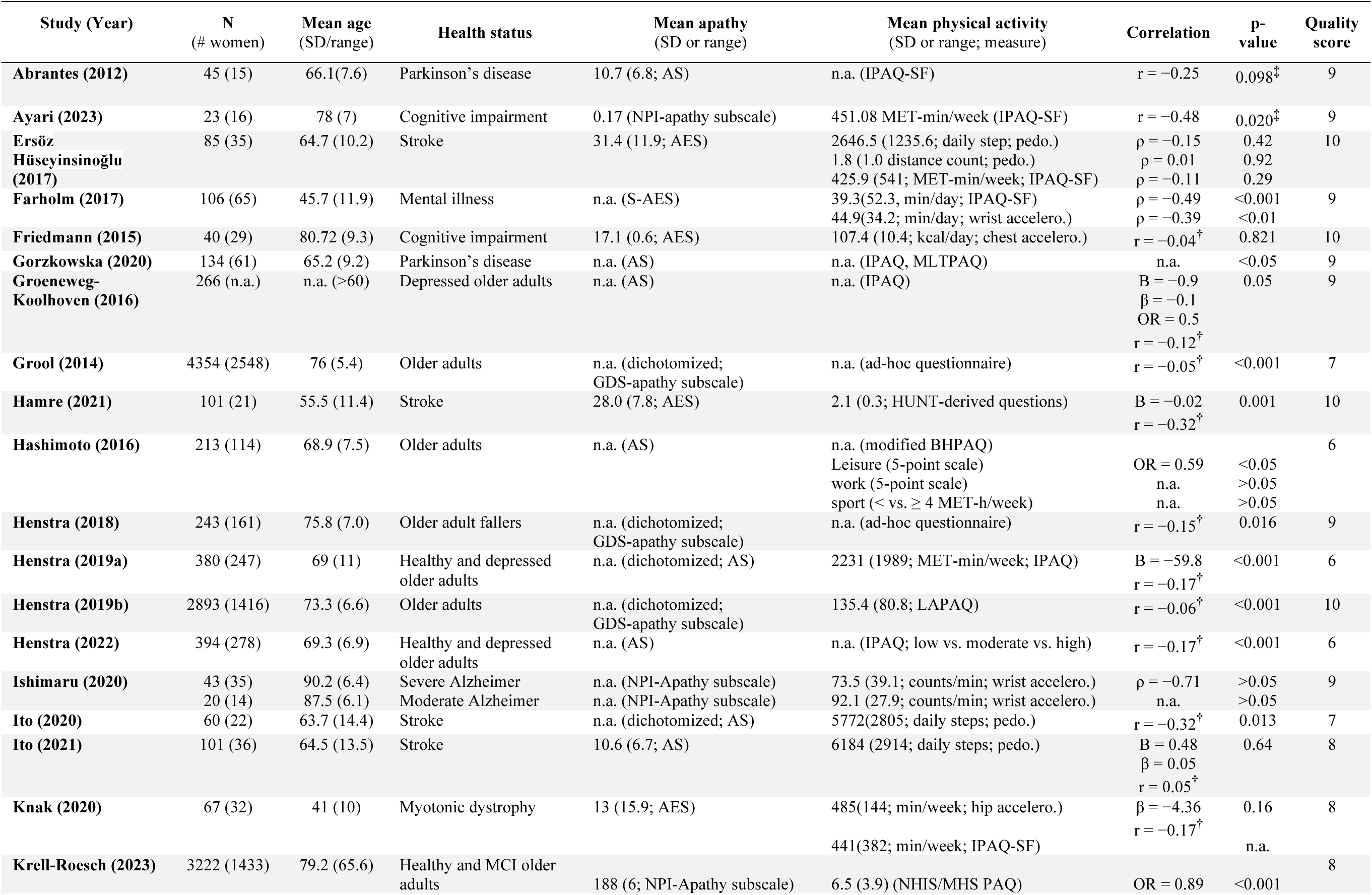
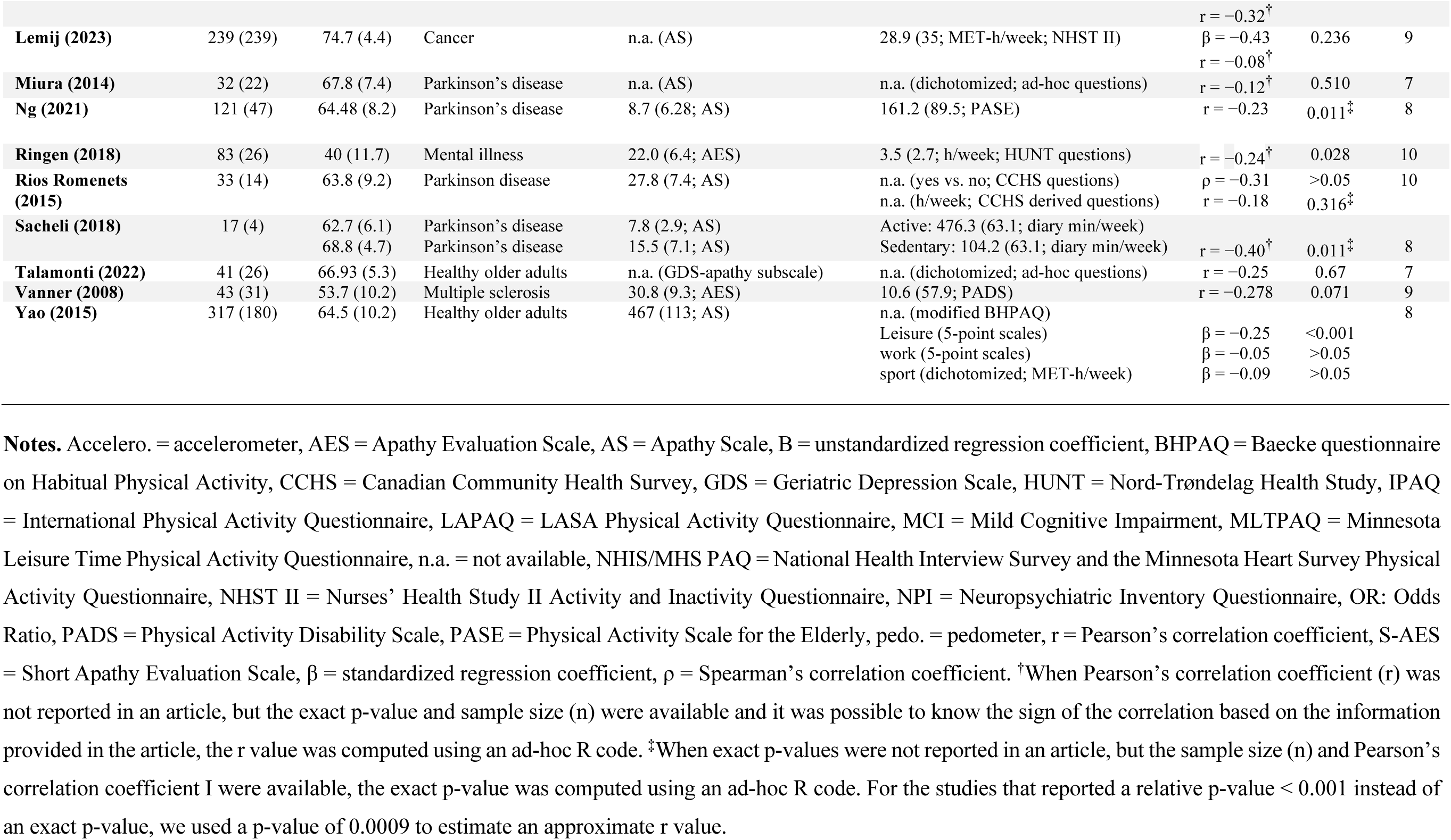
Characteristics of the studies included in the systematic review.

### 3.2. Descriptive Results

#### 3.2.1. Participants

The 28 articles identified by the systematic review included a total of 13,716 participants aged 30 to 95 years, including 7,167 women, 6,283 men, and 266 participants whose gender and sex was not reported. The studies investigated populations with stroke (k = 4) (52,53,87,88), multiple sclerosis (k = 1) (3), Parkinson’s disease (k = 6) (40,84,86,89–91), mental illness (k = 2) (92,93), cancer (k = 1) (94), myotonic dystrophy (k = 1) (51), Alzheimer’s disease (k = 1) (39), depression (k = 3) (85,95,96), cognitive impairment (k = 3) (78,79,97), as well as healthy older adults (k = 9) (38,95–102) (Table 1).

#### 3.2.2. Apathy

In 14 of the 28 studies, apathy was assessed using the Apathy Scale (k = 13) (40,52,84–87,89–91,94–96,100) or its shorter version [12-item Apathy Scale (k = 1) (38)]. This scale consists of 14 questions rated on a Likert scale ranging from 0 (not at all) to 3 (a lot). The total score on the 14-item Apathy Scale ranges from 0 to 42, with higher scores indicating more severe apathy. In clinical settings, a patient with a score ≥ 16 is considered apathetic (103). Apathy was also assessed using the Apathy Evaluation Scale (k = 6) (37,51,53,79,88,93) or its shorter version [12-item Apathy Evaluation Scale (k = 1) (92)]. This scale consists of 18 items assessing the cognitive, emotional, and behavioral aspects of apathy, rated on a Likert scale ranging from 1 (not at all) to 4 (a lot). The total score ranges from 18 to 72, with higher scores indicating more severe apathy. In clinical setting, a patient with a score ≥ 37 is considered apathetic (104). The other measures that were used are the apathy subscale of the Geriatric Depression Scale (k = 4) (98,99,101,102) and the apathy subscale of the Neuropsychiatric Inventory Questionnaire (k = 3) (39,78,97). Eighteen studies reported mean levels of apathy (Table 1). The Apathy Scale-based studies revealed a range from 8.7 (40) to 27.8 (84), while the Apathy Evaluation Scale studies reported levels between 13.0 (51) and 31.4 (53).

#### 3.2.3. Physical Activity

Twenty-four studies assessed physical activity using a self-reported measure (Table 1). Five of these questionnaire-based studies used the short form of the International Physical Activity Questionnaire (IPAQ-SF) (51,53,78,89,92), which consists of 6 items assessing time spent in light (i.e., walking), moderate (e.g., carrying light loads, cycling at moderate speed, doubles tennis), and vigorous physical activity (e.g., digging, fast cycling, heavy lifting, aerobics) over the last 7 days (105). The other questionnaires used to assess physical activity were the long form version of the IPAQ (IPAQ-LF) (k = 4) (85,86,95,96), the modified Baecke Habitual Physical Activity Questionnaire (BHPAQ) (k = 2) (38,100), the Minnesota Leisure Time Physical Activity Questionnaire (k = 1) (86), the Longitudinal Aging Study Amsterdam Physical Activity Questionnaire (LAPAQ) (k = 1) (99), the National Health Interview Survey and the Minnesota Heart Survey Physical Activity Questionnaire (NHIS/MHS PAQ) (k = 1) (97), the Nurses’ Health Study II Activity and Inactivity Questionnaire (NHST II Activity) (k = 1) (94), the Physical Activity Disability Scale (PADS) (k = 1) (37), the Nord-Trøndelag Health Study-derived questions (HUNT) (k = 2) (88,93), the Canadian Community Health Survey-derived questionnaire (CCHS) (k = 1) (84), and the Physical Activity Scale for the Elderly (PASE) (k = 1) (40).

Physical activity was also assessed with devices such as accelerometers measuring accelerations in three dimensions (k = 4) (39,51,79,92) and pedometers measuring the number of steps (k = 3) (52,53,87) (Table 1). These devices were worn at the hip (k = 2) (51,53), chest (k = 1) (79), wrist (k = 2) (39,92), or pocket (k = 1) (52), and one study did not report where the device was worn (87). Studies based on accelerometer-derived measures used an ActiGraph device (ActiGraph, LLC, Pensacola, FL, USA) (51,79), the Polar M200 (Polar Electro Oy, Kempele, Finland) (92), or the Micro Motionlogger Watch (Ambulatory Monitoring, Ardsley, NY, USA) (39). The pedometers used were the Omron Walking Style Pro 2.0 (Omron Healthcare, Kyoto, Japan) (53), or the Yamasa EX-300 (Yamasa, Choshi, Japan) (52). One study did not specify the pedometer that was used (87). To assess physical activity, the studies used the following outcomes: Score from a questionnaire (e.g., PASE, PADS, BHPAQ) (k = 11) (37,38,40,84,88,91,97–101), METmin/week (k = 7) (38,53,85,94–96,100), steps per day (k = 3) (52,53,87), active time per day or week (k = 8) (51,84,86,89,90,92,93,102), counts per minute (k = 1) (39), kilocalories per day (k = 1) (79). Four studies used multiple physical activity outcomes (k = 4) (38,53,84,100).

#### 3.2.4. Association Between Physical Activity and Apathy

Among the 28 articles included in the systematic review, five reported correlation coefficients of the association between physical activity and apathy. Specifically, two articles reported at least one Pearson’s r correlation coefficient (37,89) and three other articles reported at least one Spearman’s rho (39,53,92). The Pearson’s r value was computed in 11 studies (51,52,79,85,87,88,90,91,93,94,98) using the exact p value (or t value) and sample size (Supplemental R Script 1). An approximate r value was estimated for five studies reporting a relative p-value < 0.001 (95–97,99,101). Through email correspondence with the authors, we obtained three additional Pearson’s r values (40,78,84) and one additional Spearman’s rho value (85). Furthermore, one Pearson’s r value was calculated based on publicly available data (102). In total, 22 Pearson’s r values from 22 studies (37,39,51,52,78,79,84,85,87–91,93–101) and seven Spearman’s rho values from four studies (39,53,84,92) were used in the meta-analysis (Table 1). The remaining 3 studies did not report a correlation coefficient and were therefore not included in the meta-analysis. Two of these studies reported a significant negative association between physical activity and apathy based on odds ratios (100) or standardized beta coefficient (β) (38). The third study reported only a relative p-value > 0.05 (86).

### 3.3. Meta-Analysis

#### 3.3.1. Main meta-analysis

Our meta-analysis of 22 studies (n = 12,541) based on Pearson’s r values revealed a statistically significant small negative correlation between apathy and physical activity (r = -0.13; 95% confidence interval [95% CI]: -0.18 to -0.09; p < 0.0001; Table 2; Figure 2). Further supporting this result, between-study statistical heterogeneity could be considered moderate (Tau^2^ = 0.0038; 95% CI: 0.0003 to 0.0167; I^2^ = 49.0%; 95% CI: 16.4 to 68.9%), and the prediction interval ranged from r = -0.26 to 0.00, suggesting that the correlation is expected to be negative for a future study.

**Table 2.**
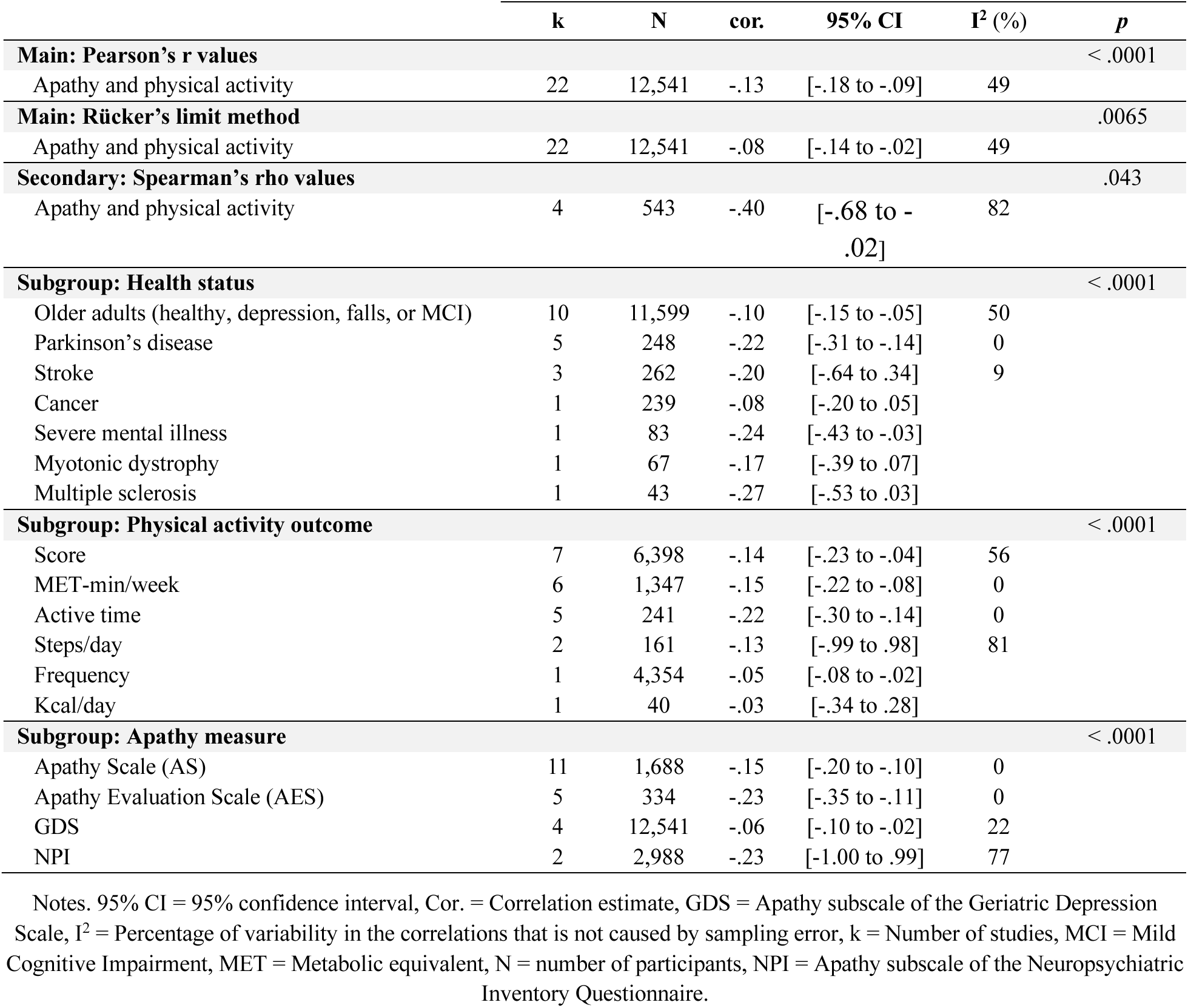
Results of the main, secondary, and subgroup meta-analyses

**Figure 2.**
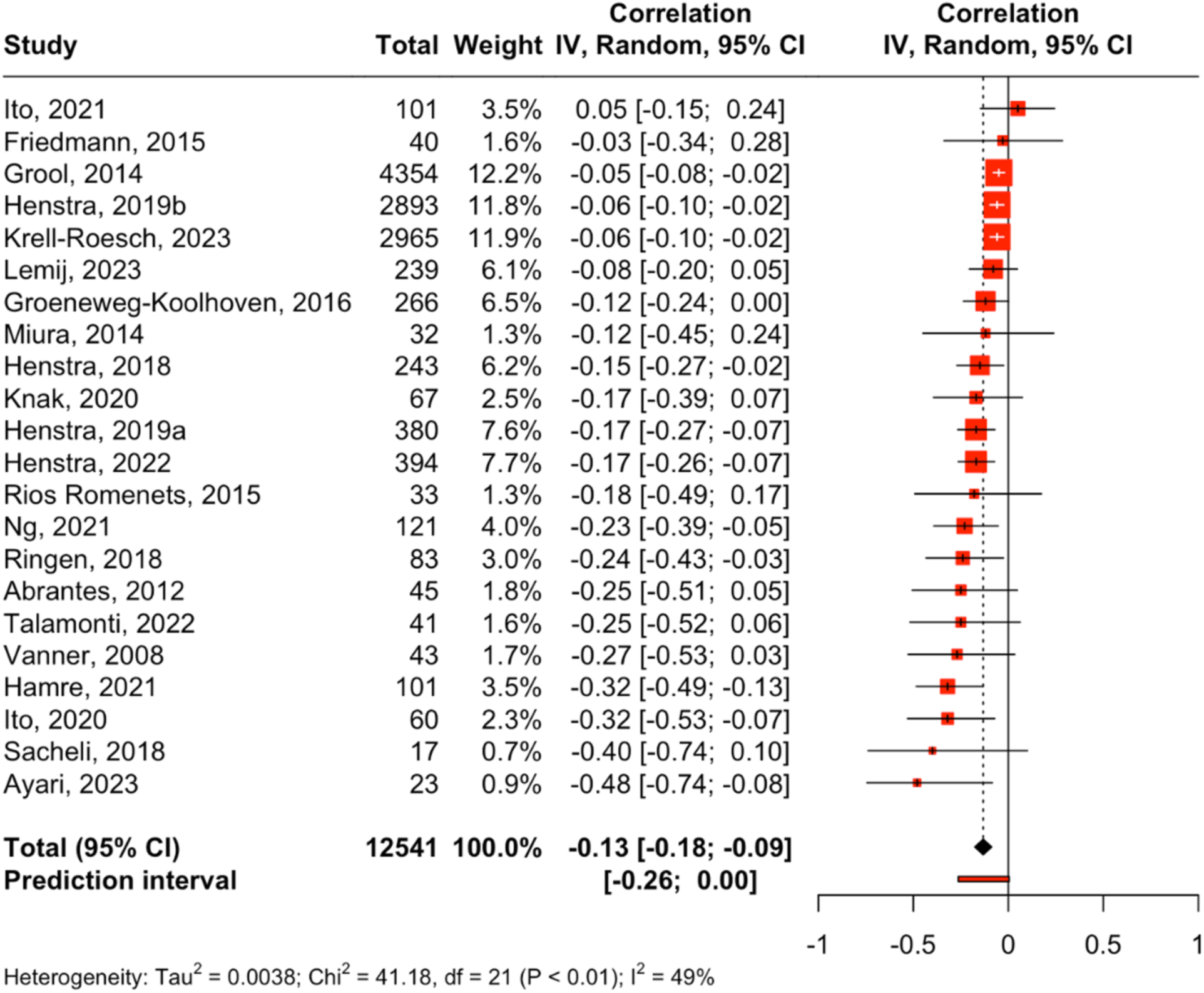
Main meta-analysis. Correlation between apathy and physical activity based on Pearson’s r values (k = 22, n = 12,541). CI = confidence interval, IV = inverse variance.

#### 3.3.2. Publication Bias Assessment

The funnel plot showed an asymmetrical pattern (Figure 3A), with more studies on the left of the vertical dashed line representing the average effect size. In addition, if the missing studies were imputed in the right part of the plot to increase the symmetry, most of these studies would lie in the non-significance region (in white), which suggests that the asymmetry in the funnel plot may be caused by publication bias rather than other potential causes, such as different study procedures and between-study heterogeneity. Egger’s regression test confirmed that the data in the funnel plot was asymmetric (b = -1.44; 95% CI: -1.97 to -0.91; p = 3.1 × 10^-5^). However, the bias-corrected estimate of the true effect size, calculated using Rücker’s limit meta-analysis method, showed that the small correlation reported in the main analysis would remain significant even if such publication bias was present in our data set (r = -0.08; 95% CI: -0.14 to -0.02; p = 0.0065).

**Figure 3.**
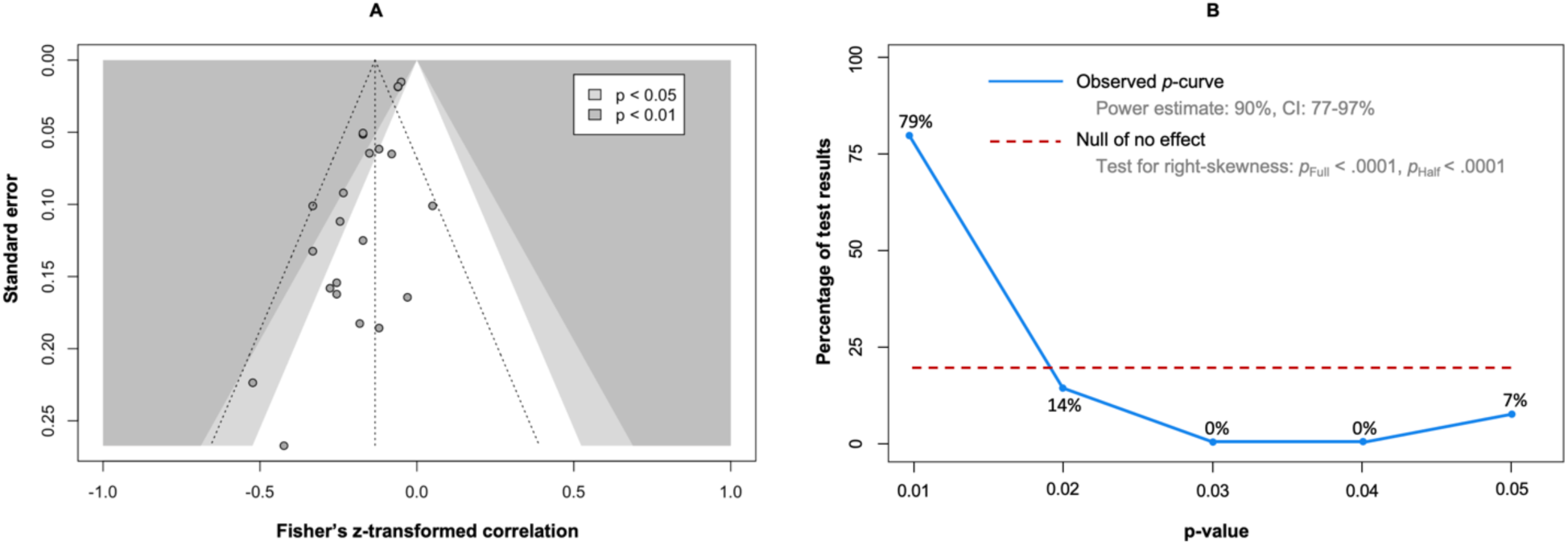
Publication bias assessment. Contour-enhanced funnel plot of the main meta-analysis (A) The vertical dashed line represents the average effect size. The two other dashed lines represent the idealized funnel-shape that studies are expected to follow. When there is no publication bias, the data points in a funnel plot should form a roughly symmetrical, upside-down funnel. Studies in the top part of the plot, which have lower standard errors, are expected to lie closely together, and not far away from the pooled effect size. In the lower part of the plot, studies have higher standard errors, the funnel “opens up”, and effect sizes are expected to scatter more heavily to the left and right of the pooled effect. **P-curve analysis (B).** The blue line indicates the distribution of the analyzed p-values. The red dotted line illustrates a uniform distribution of the p-values, indicating the absence of a true effect.

The 29 correlation values (Pearson’s r or Spearman’s rho) were provided to the p-curve analysis (Figure 3B). The observed p-curve included 14 statistically significant results (p < 0.05), 13 of which were highly significant (p < 0.025). The p-value of the right-skewness test was < 0.001 for both the half curve (curve of p values ≤ 0.025) and the full curve (curve of p values < 0.05), confirming that the p-curve was right-skewed and suggesting that the effect of our meta-analysis is true, i.e., that the effect we estimated is not an artifact caused by selective reporting (e.g., p-hacking) in the literature (106). In addition, the statistical power of the studies that were included in the p-curve analysis was 90% (90% CI: 77 to 97%), suggesting that approximately 90% of the significant results are expected to be replicable.

#### 3.3.3. Secondary Meta-Analysis

Results of the secondary meta-analysis based on Spearman’s rho values (k = 4, n = 543) were consistent with those based on Pearson’s r as they showed a statistically significant moderate to large negative correlation between apathy and physical activity (r = -0.40; 95% CI: -0.68 to -0.02; p = 0.043) (Figure 4). However, we observed substantial to considerable between-study statistical heterogeneity (between-cluster Tau^2^ = 0.09; 95% CI: 0.01 to 0.97; I^2^ = 82.0%; 95% CI: 64.1 to 91.0%), and the prediction interval ranged from r = -0.87 to 0.45, indicating that a moderate to large positive correlation cannot be ruled out for future studies.

**Figure 4.**
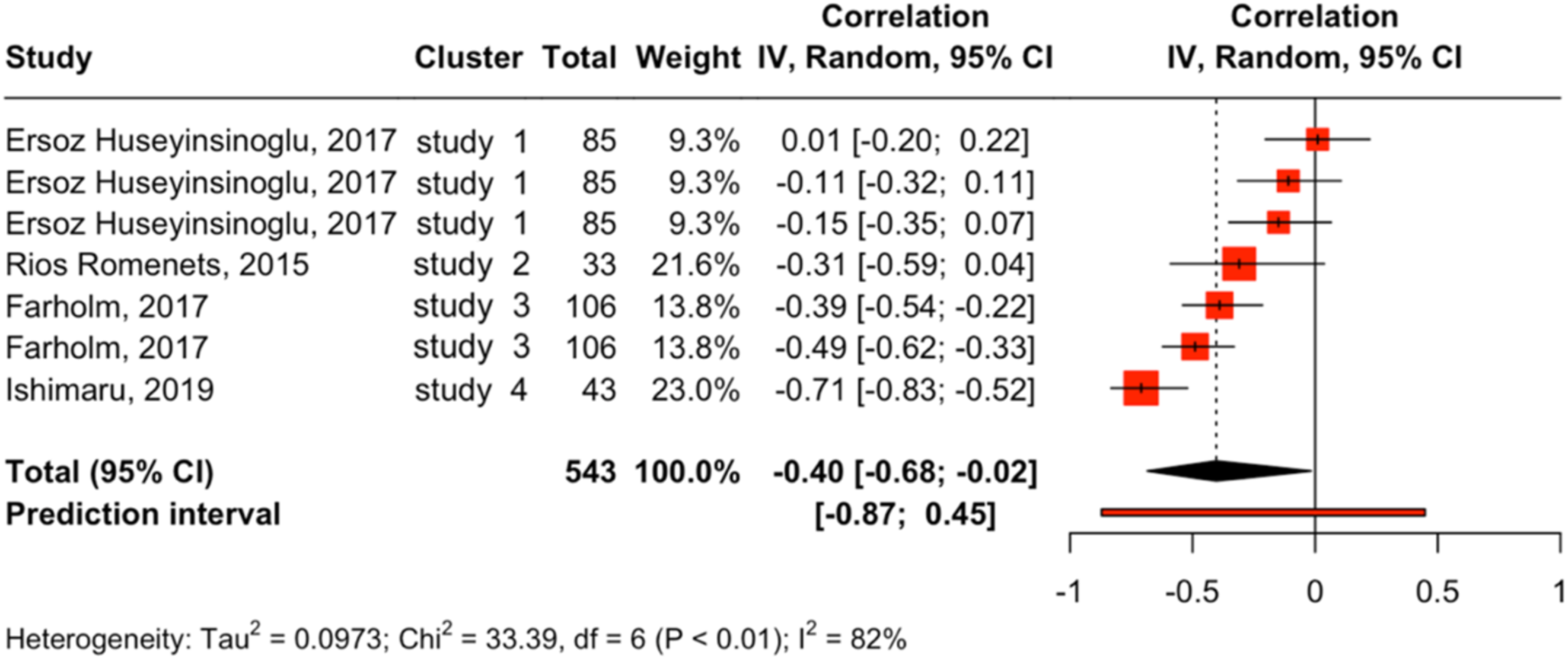
Secondary meta-analysis. Correlation between apathy and physical activity based on Spearman’s rho values (k = 7, n = 437). CI = confidence interval, IV = inverse variance.

The sampling error variance on level 1 and the value of I^2^ on level 2 (i.e., the amount of heterogeneity variance within studies) were small (12.4% and 3.5 × 10^-9^ %, respectively). The largest share of heterogeneity variance was from level 3, with between-study heterogeneity making up 87.6% of the total variation in our data. Overall, this indicates that there is considerable between-study heterogeneity, and that little variance can be explained by differences within studies. However, the 3-level model showed a better fit than the 2-level model with lower Akaike’s information criterion (AIC) (3.2 vs. 6.6) and Bayesian information criterion (BIC) (2.6 vs. 6.2), indicating better performance. These lower AIC and BIC are consistent with the significant likelihood ratio test (LRT) comparing the two models (χ^2^ = 5.35; p = 0.0207). Therefore, although the 3-level model introduces an additional parameter, this added complexity has improved our estimate of the pooled effect.

#### 3.3.4. Subgroup Meta-Analyses

To test for subgroup differences between health status, we compared studies comprising older adults who were healthy, depressed, fallers, or had mild cognitive impairment (k = 10), people with Parkinson’s Disease (k = 5), and stroke survivors (k = 3). We found statistical difference between these studies (p < 0.0001) (Table 2; Supplemental Figure 1). The relationship between apathy and physical activity was statistically significant in studies that included older adults (r = - 0.10; 95% CI: -0.15 to -0.05) or patients with Parkinson’s disease (r = -0.22; 95% CI: -0.31 to - 0.14), but not in studies that included stroke survivors (r = -0.20; 95% CI: -0.64 to -0.34). However, statistical power was lacking in the latter (k = 3) and other health status (k = 1).

To test for subgroup differences between physical activity outcomes, we compared studies using a score from a questionnaire (k = 7), MET-min per week (k = 6), active time per day or week (k = 5), and steps per day (k = 2) (Table 2; Supplemental Figure 2). We found statistical difference between these studies (p < 0.0001). The relationship between apathy and physical activity was statistically significant in studies using a score (r = -0.14; 95% CI: -0.23 to -0.04), MET-min/week (r = -0.15; 95% CI: -0.22 to -0.08), and active time (r = -0.22; 95% CI: -0.30 to -0.14), but not in studies that used the number of steps per day (r = -0.13; 95% CI: -0.99 to 0.98). However, statistical power was lacking in the latter (k = 2) and other physical activity outcomes (k = 1).

To test for subgroup differences between apathy measures, we compared studies using the Apathy Scale (k = 11), the Apathy Evaluation Scale (k = 5), the apathy subscale of the Geriatric Depression Scale (k = 4), and the apathy subscale of the Neuropsychiatric Inventory Questionnaire (k = 2) (Table 2; Supplemental Figure 3). The relationship between apathy and physical activity was statistically significant in studies using the Apathy Scale (r = -0.15; 95% CI: -0.20 to -0.10), Apathy Evaluation Scale (r = -0.23; 95% CI: -0.35 to -0.11), and Geriatric Depression Scale (r = - 0.06; 95% CI: -0.10 to -0.02), but not in studies that used the Neuropsychiatric Inventory Questionnaire (r = -0.23; 95% CI: -1.00 to 0.99). However, statistical power was lacking in the latter apathy measure (k = 2).

#### 3.3.5. Meta-Regressions

Age statistically influenced the correlation values of the meta-analysis studies (k = 21; p = 0.003) (Figure 5A), with older samples being associated with more negative relationships between apathy and physical activity. Conversely, the proportion of women did not statistically influence the meta-analysis studies’ correlation values (k = 21; p = 0.346) (Figure 5B).

**Figure 5.**
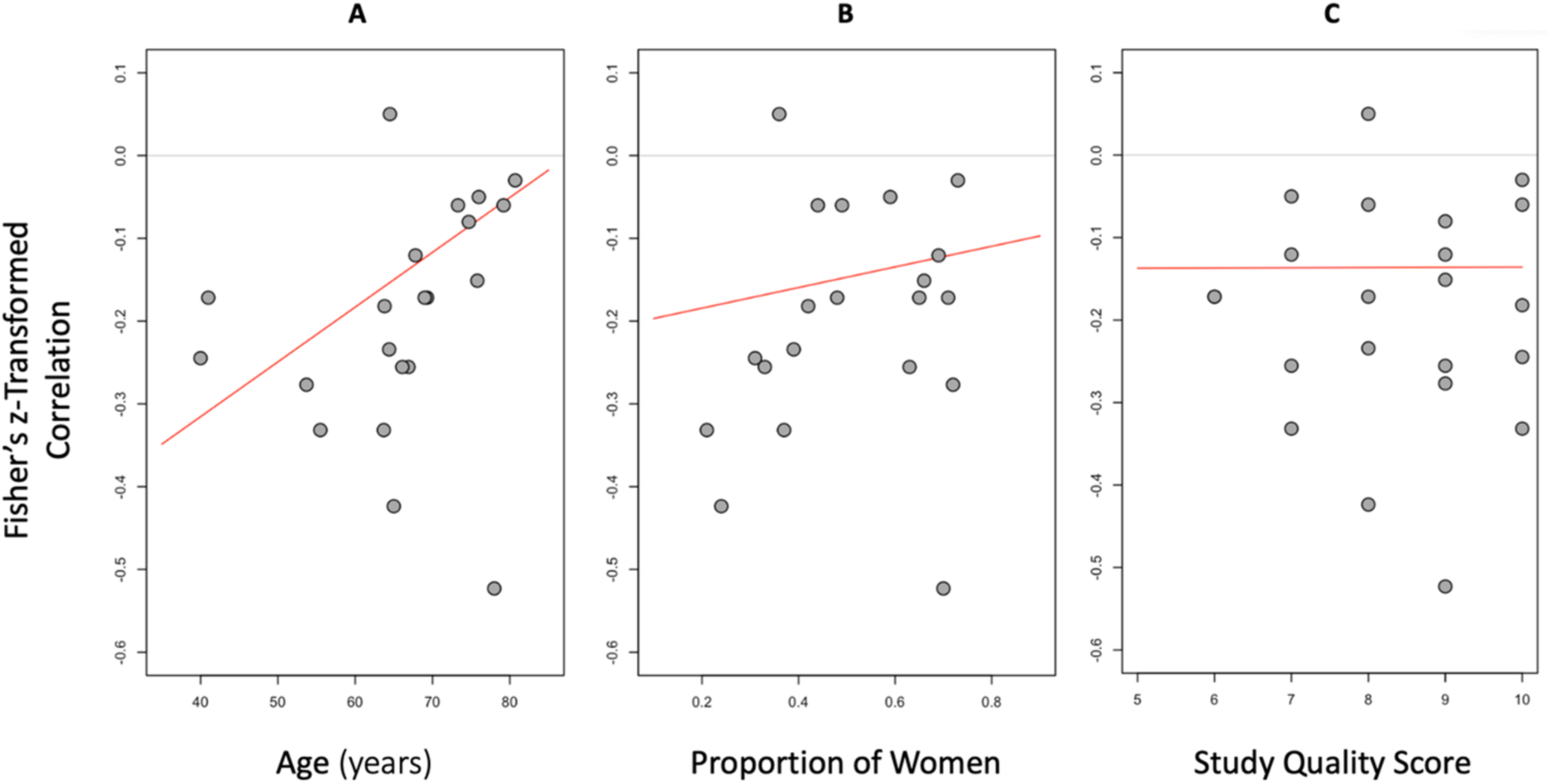
Meta-Regressions testing the influence of age (A), the proportion of women (B), and study quality score (C) on the correlation estimates of the main meta-analysis studies.

#### 3.3.6. Sensitivity Analysis

The meta-regression by quality score based on the Quality Assessment Tool for Observational Cohort and Cross-Sectional Studies (61) (k = 22) showed that a study’s quality did not influence correlation values (p = 0.986) (Figure 5C).

## 4. Discussion

### 4.1. Clinical Implications

As hypothesized, the main meta-analysis based on Pearson’s r values showed a small negative correlation between apathy and physical activity. The secondary analysis based on Spearman’s rho values showed a moderate to large negative correlation, further supporting the results. While larger effect sizes may result in more noticeable clinical differences at the individual level and in the short term, smaller effect sizes may also have important clinical implications, especially when considered in the context of population health, prevention strategies, and long-term outcomes. Given the important contribution of physical activity to the primary, secondary, and tertiary prevention of many diseases including cardiovascular disease (107,108), stroke (49,109), depression (110,111), obesity (112), dementia (113), osteoarthritis (114), and cancer (115), the small effect size we report in the main meta-analysis based on Pearson’s r values may translate into meaningful clinical improvements in patient outcomes.

Health professionals have an important role to play in preventing and reducing the detrimental effect of apathy on physical activity. Non-pharmacological interventions such as multisensory stimulation, music therapy, cognitive stimulation, and pet therapy have been shown to be effective in decreasing apathy (114). Physical therapists can contribute through exercise therapy as intervention studies have shown that exercise can reduce apathy (115,116), opening the possibility for a virtuous circle as this reduced apathy can then favor physical activity engagement.

In patients with apathy, the exercise intervention should take into consideration the reduced motivation, as this will affect the patient’s ability to adhere to the prescribed program (3). Since the perceived cost of physical activity is higher in this population, the therapist needs to find ways to increase the rewards associated with the intervention. These rewards can be intrinsic (e.g., fun activity) or extrinsic (e.g., validation of the progression checks set by the therapist).

### 4.2. Underlying Mechanisms and Health Status

The mechanisms underlying the association between apathy and physical activity remain poorly understood and overlooked. One exception is a recent study showing that weaker intention to be physically active mediated the association between higher levels of behavioral apathy and lower habitual levels of moderate-to-vigorous physical activity (119). In addition, explicit attitudes mediated the effect of behavioral apathy on intentions to be physically active. These findings indicate that controlled motivational processes are involved, allowing for potential interventions to reduce the detrimental effects of apathy on physical activity.

In our meta-analysis, the largest significant effect size was observed in patients with Parkinson’s disease, which can be used as a model to better understand the mechanisms underlying the association between apathy and physical activity. This association is consistent with the decision theory of goal-directed behavior (120). According to this theory, people contrast costs and benefits when considering whether to engage in physical activity. Engagement occurs when the net value of this contrast is beneficial. In patients with Parkinson’s disease, apathy is thought to result from dopamine depletion in the brain (121–125). When this depletion is treated with dopaminergic medications, studies have shown that patients are more willing to squeeze a handgrip to earn a reward than to decline (126), and they exert more force to earn a reward (127,128). Taken together, these results suggest that in patients with Parkinson’s disease, apathy may be associated with dopaminergic deficits that could increase the perceived cost of physical effort, thereby decreasing the willingness to engage in physical activity.

In addition to patients with Parkinson’s disease, our subgroup meta-analyses suggests that the negative relationship between apathy and physical activity was observed in older adults who were healthy, depressed, fallers, or had mild cognitive impairment. This result was further supported by the meta-regression showing that older ages were associated with more negative relationships between apathy and physical activity. This effect of aging could be explained that the higher metabolic energy associated with a given physical activity in older compared to young adults (129–131). This decreased efficiency may increase the cost for engaging in physical activity in a non-linear manner across the years, amplifying the impact of apathy on the engagement in physical activity. Moreover, apathy is thought to increase with age (55), which would further contribute to amplify the impact of this decreased efficiency on physical activity.

Although there was no clear evidence of an association between apathy and physical activity in the other health conditions, such an association cannot be ruled out, as the lack of statistical significance could be explained by a lack of statistical power in these health conditions (e.g., stroke) or by the impossibility of comparing them with other health conditions in the subgroup meta-analysis, as they were only examined in a single study (e.g., cancer). Contrary to previous studies (55,56,132), our results showed no evidence of an effect of gender.

### 4.3. Measures of Physical Activity and Apathy

Both self-reported (e.g., MET-minutes per week) and device-based outcomes (e.g., active time per day or per week) of physical activity were negatively associated with apathy, suggesting that both these approaches can capture this relationship. The number of steps per day was not associated with apathy, but this analysis may be underpowered because it included only two studies with a total of 161 participants, some of whom may have been included in these two studies from the same group with similar methods and partly overlapping recruitment periods (52,87).

The three most used measures of apathy (Apathy Scale, Apathy Evaluation Scale, Apathy Subscale of the Geriatric Depression Scale) were all associated with physical activity, further supporting the robustness of this association. Only one measure of apathy showed no evidence of an association with physical activity, but this result was based on only two studies, which both showed a significant negative association.

### 4.4. Limitations

The results of this systematic review and meta-analysis should be considered in light of several limitations. (i) Only articles published in English were included. Inclusion of articles published in other languages may have influenced the results. (ii) Due to the correlational nature of the meta-analysed results, we cannot conclude a causal relationship between apathy and physical activity, nor can we exclude the possibility that this relationship is indirect, i.e., mediated by other factors. (iii) The meta-analysis was not pre-registered because the author with this expertise (IML) joined only after the systematic review was conducted. However, since the pre-registered systematic review defines the studies included in the meta-analysis and since the ‘meta’ R package (63) provides very few degrees of freedom, there was little room for bias in the main and secondary meta-analyses. This reasoning also applies for the subgroup analyses as the health conditions, types of physical activity measures, measurement instruments, and outcomes, as well as the apathy measurement instruments, were imposed by the studies that were included in the systematic review. (iv) Only 3 of the 43 authors we contacted (7%) shared their estimates (n = 2) or raw data (n = 1) with us, which is less than reported in previous literature (133). Including these estimates may have influenced our results.

## 5. Conclusion

Our results suggest that higher levels of apathy are associated with lower levels of physical activity and that this negative association is stronger with age. As such, apathy may be a limiting factor to exercise therapy, which is within the scope of practice of physiotherapists (134) and may have prognostic implications in patients whose condition requires physical activity. Accordingly, physiotherapists should screen for apathy when a patient has difficulty engaging in physical activity, especially older adults, and develop strategies to increase the intrinsic and extrinsic motivation associated with physical activity in those who experience apathy.

## 6. Declarations

### 6.1. Data and Code Availability

According to good research practices (135), the dataset as wells as R and R Markdown scripts are freely available in Zenodo (136).

### 6.2. Authorship Contribution Statement

Based on the Contributor Roles Taxonomy (CRediT) (137) individual author contributions to this work are as follows:

- Ata Farajzadeh: Conceptualization; Methodology (systematic review); Formal Analysis; Investigation; Writing – Original Draft; Writing – Review and Editing.
- Alexe Hébert: Investigation.
- Ian M. Lahart: Methodology (meta-analysis); Formal Analysis; Writing – Review and Editing.
- Martin Bilodeau: Writing – Review and Editing; Supervision of AH.
- Matthieu P. Boisgontier: Conceptualization; Methodology; Formal Analysis; Data Curation; Visualization; Writing – Original Draft; Writing – Review and Editing; Supervision of AF and AH; Project Administration; Funding Acquisition.

## Data Availability

All data produced are available online at https://doi.org/10.5281/zenodo.10929857

## SUPPLEMENTAL MATERIAL

**Supplemental Material 1.** Search strategies for the PubMed, PsycINFO, Web of Science, SPORTDiscus, CINHAL, and Embase databases

**Supplemental Material 2.** Model underlying Rücker’s limit meta-analysis method.

**Supplemental R Script 1**. R script for the calculation of Pearson’s r estimate based on the degrees of freedom (sample size - 2) and exact p-value, when the direction of the relationship is known.

**Supplemental R Script 2**. R script for the calculation of exact p-values based on sample size (n) and Pearson’s correlation coefficient (r).

**Supplemental Figure 1.** Subgroup meta-analyses. Differences according to health status.

**Supplemental Figure 2.** Subgroup meta-analyses. Differences according to physical activity outcome.

**Supplemental Figure 3.** Subgroup meta-analyses. Differences according to apathy measure.

**Supplemental Material 1.** Search strategies for the PubMed, PsycINFO, Web of Science, SPORTDiscus, CINHAL, and Embase databases

### PubMed

(((((((((((((((((((((“apathy”[Title/Abstract]) OR (“apathetic”[Title/Abstract])) OR(“abulia”[Title/Abstract])) OR (“amotivation”[Title/Abstract])) OR (“avolition”[Title/Abstract])) OR (“neuropsychiatric inventory”[Title/Abstract])) OR (“NPI”[Title/Abstract])) OR (“emotional indifference”[Title/Abstract])) OR (“frontal lobe personality scale”[Title/Abstract])) OR (“Lille apathy rating scale”[Title/Abstract])) OR (“LARS”[Title/Abstract])) OR (“dementia apathy interview and rating”[Title/Abstract])) OR (“DAIR”[Title/Abstract])) OR (“frontal system behavior scale”[Title/Abstract])) OR (“FrSBe”[Title/Abstract])) OR (“key behaviors change inventory”[Title/Abstract])) OR (“KBCI”[Title/Abstract])) OR (“apathy evaluation scale”[Title/Abstract])) OR (“apathy scale”[Title/Abstract])) OR (“irritability apathy scale”[Title/Abstract])) OR (“IAS”[Title/Abstract])) AND (((((((“physical activity”[Title/Abstract]) OR (“physical education”[Title/Abstract])) OR (“training”[Title/Abstract])) OR (“physical fitness”[Title/Abstract])) OR (“exercise”[Title/Abstract])) OR (“sport”[Title/Abstract])) OR (“walk”[Title/Abstract]))

### PsycINFO

1. Physical Activity/
2. physical education.mp. [mp=title, abstract, heading word, table of contents, key concepts, original title, tests & measures, mesh word]
3. training.mp. [mp=title, abstract, heading word, table of contents, key concepts, original title, tests & measures, mesh word]
4. physical fitness.mp. [mp=title, abstract, heading word, table of contents, key concepts, original title, tests & measures, mesh word]
5. exercise.mp. [mp=title, abstract, heading word, table of contents, key concepts, original title, tests & measures, mesh word]
6. sport.mp. [mp=title, abstract, heading word, table of contents, key concepts, original title, tests & measures, mesh word]
7. walk.mp. [mp=title, abstract, heading word, table of contents, key concepts, original title, tests & measures, mesh word]
8. Apathy/
9. abulia.mp. [mp=title, abstract, heading word, table of contents, key concepts, original title, tests & measures, mesh word]
10. apathetic.mp. [mp=title, abstract, heading word, table of contents, key concepts, original title, tests & measures, mesh word]
11. amotivation.mp. [mp=title, abstract, heading word, table of contents, key concepts, original title, tests & measures, mesh word]
12. avolition.mp. [mp=title, abstract, heading word, table of contents, key concepts, original title, tests & measures, mesh word]
13. neuropsychiatric inventory.mp. [mp=title, abstract, heading word, table of contents, key concepts, original title, tests & measures, mesh word]
14. NPI.mp. [mp=title, abstract, heading word, table of contents, key concepts, original title, tests & measures, mesh word]
15. emotional indifference.mp. [mp=title, abstract, heading word, table of contents, key concepts, original title, tests & measures, mesh word]
16. frontal lobe personality scale.mp. [mp=title, abstract, heading word, table of contents, key concepts, original title, tests & measures, mesh word]
17. Lille apathy rating scale.mp. [mp=title, abstract, heading word, table of contents, key concepts, original title, tests & measures, mesh word]
18. LARS.mp. [mp=title, abstract, heading word, table of contents, key concepts, original title, tests & measures, mesh word]
19. dementia apathy interview and rating.mp. [mp=title, abstract, heading word, table of contents, key concepts, original title, tests & measures, mesh word]
20. DAIR.mp. [mp=title, abstract, heading word, table of contents, key concepts, original title, tests & measures, mesh word]
21. frontal system behavior scale.mp. [mp=title, abstract, heading word, table of contents, key concepts, original title, tests & measures, mesh word]
22. FrSBe.mp. [mp=title, abstract, heading word, table of contents, key concepts, original title, tests & measures, mesh word]
23. key behaviors change inventory.mp. [mp=title, abstract, heading word, table of contents, key concepts, original title, tests & measures, mesh word]
24. KBCI.mp. [mp=title, abstract, heading word, table of contents, key concepts, original title, tests & measures, mesh word]
25. apathy evaluation scale.mp. [mp=title, abstract, heading word, table of contents, key concepts, original title, tests & measures, mesh word]
26. apathy scale.mp. [mp=title, abstract, heading word, table of contents, key concepts, original title, tests & measures, mesh word]
27. irritability apathy scale.mp. [mp=title, abstract, heading word, table of contents, key concepts, original title, tests & measures, mesh word]
28. IAS.mp. [mp=title, abstract, heading word, table of contents, key concepts, original title, tests & measures, mesh word]
29. 1 or 2 or 3 or 4 or 5 or 6 or 7
30. 8 or 9 or 10 or 11 or 12 or 13 or 14 or 15 or 16 or 17 or 18 or 19 or 20 or 21 or 22 or 23 or 24 or 25 or 26 or 27 or 28
31. 29 and 30

### Web of Science

1: ((((((((AB=(Physical activity)) OR AB=(physical education)) OR AB=(training)) OR AB=(physical fitness)) OR AB=(exercise)) OR AB=(sport)) OR AB=(walk)))

2: ((((((((((((((((((((AB=(apathy)) OR AB=(abulia)) OR AB=(apathetic)) OR AB=(amotivation)) OR AB=(avolition)) OR AB=(neuropsychiatric inventory)) OR AB=(NPI)) OR AB=(emotional indifference)) OR AB=(frontal lobe personality scale)) OR AB=(Lille apathy rating scale)) OR AB=(LARS)) OR AB=(dementia apathy interview and rating)) OR AB=(DAIR)) OR AB=(frontal system behavior scale)) OR AB=(FrSBe)) OR AB=(key behaviors change inventory)) OR AB=(KBCI)) OR AB=(apathy evaluation scale)) OR AB=(apathy scale)) OR AB=(irritability apathy scale)) OR AB=(IAS)

3: #2 AND #1

### SPORTDiscus

S1: AB physical activity OR AB physical education OR AB walk OR AB sport OR AB exercise OR AB physical fitness OR AB training

S2: AB apathy OR AB abulia OR AB apathetic OR AB amotivation OR AB avolition OR AB neuropsychiatric inventory OR AB NPI OR AB emotional indifference OR AB frontal lobe personality scale OR AB Lille apathy rating scale OR AB LARS OR AB dementia apathy interview and rating OR AB DAIR OR AB frontal system behavior scale OR AB FrSBe OR AB key behaviors change inventory OR AB KBCI OR AB apathy evaluation scale OR AB apathy scale OR AB irritability apathy scale OR AB IAS

S3: S1 AND S2

### CINAHL

S1: AB physical activity OR AB physical education OR AB exercise OR AB physical fitness OR AB sport OR AB training OR AB walk

S2: AB apathy OR AB abulia OR AB amotivation OR AB apathetic OR AB avolition OR AB emotional indifference OR AB neuropsychiatric inventory OR AB Lille apathy rating scale OR AB NPI OR AB LARS OR AB frontal system behavior scale OR AB FrSBe OR AB key behaviors change inventory OR AB KBCI OR AB apathy evaluation scale OR AB apathy scale OR AB irritability apathy scale OR AB IAS OR AB dementia apathy interview and rating OR AB DARS

S3: S1 AND S2

### Embase

1. physical activity/
2. physical education.mp. [mp=title, abstract, heading word, drug trade name, original title, device manufacturer, drug manufacturer, device trade name, keyword heading word, floating subheading word, candidate term word]
3. exercise.mp. [mp=title, abstract, heading word, drug trade name, original title, device manufacturer, drug manufacturer, device trade name, keyword heading word, floating subheading word, candidate term word]
4. training.mp. [mp=title, abstract, heading word, drug trade name, original title, device manufacturer, drug manufacturer, device trade name, keyword heading word, floating subheading word, candidate term word]
5. walk.mp. [mp=title, abstract, heading word, drug trade name, original title, device manufacturer, drug manufacturer, device trade name, keyword heading word, floating subheading word, candidate term word]
6. physical fitness.mp. [mp=title, abstract, heading word, drug trade name, original title, device manufacturer, drug manufacturer, device trade name, keyword heading word, floating subheading word, candidate term word]
7. sport.mp. [mp=title, abstract, heading word, drug trade name, original title, device manufacturer, drug manufacturer, device trade name, keyword heading word, floating subheading word, candidate term word]
8. 1 or 2 or 3 or 4 or 5 or 6 or 7
9. apathy/
10. abulia.mp. [mp=title, abstract, heading word, drug trade name, original title, device manufacturer, drug manufacturer, device trade name, keyword heading word, floating subheading word, candidate term word]
11. amotivation.mp. [mp=title, abstract, heading word, drug trade name, original title, device manufacturer, drug
12. apathetic.mp. [mp=title, abstract, heading word, drug trade name, original title, device manufacturer, drug manufacturer, device trade name, keyword heading word, floating subheading word, candidate term word]
13. avolition.mp. [mp=title, abstract, heading word, drug trade name, original title, device manufacturer, drug manufacturer, device trade name, keyword heading word, floating subheading word, candidate term word]
14. neuropsychiatric inventory.mp. [mp=title, abstract, heading word, drug trade name, original title, device manufacturer, drug manufacturer, device trade name, keyword heading word, floating subheading word, candidate term word]
15. NPI.mp. [mp=title, abstract, heading word, drug trade name, original title, device manufacturer, drug manufacturer, device trade name, keyword heading word, floating subheading word, candidate term word]
16. emotional indifference.mp. [mp=title, abstract, heading word, drug trade name, original title, device manufacturer, drug manufacturer, device trade name, keyword heading word, floating subheading word, candidate term word]
17. frontal lobe personality scale.mp. [mp=title, abstract, heading word, drug trade name, original title, device manufacturer, drug manufacturer, device trade name, keyword heading word, floating subheading word, candidate term word]
18. Lille apathy rating scale.mp. [mp=title, abstract, heading word, drug trade name, original title, device manufacturer, drug manufacturer, device trade name, keyword heading word, floating subheading word, candidate term word]
19. LARS.mp. [mp=title, abstract, heading word, drug trade name, original title, device manufacturer, drug manufacturer, device trade name, keyword heading word, floating subheading word, candidate term word]
20. dementia apathy interview and rating.mp. [mp=title, abstract, heading word, drug trade name, original title, device manufacturer, drug manufacturer, device trade name, keyword heading word, floating subheading word, candidate term word]
21. DAIR.mp. [mp=title, abstract, heading word, drug trade name, original title, device manufacturer, drug manufacturer, device trade name, keyword heading word, floating subheading word, candidate term word]
22. frontal system behavior scale.mp. [mp=title, abstract, heading word, drug trade name, original title, device manufacturer, drug manufacturer, device trade name, keyword heading word, floating subheading word, candidate term word]
23. FrSBe.mp. [mp=title, abstract, heading word, drug trade name, original title, device manufacturer, drug manufacturer, device trade name, keyword heading word, floating subheading word, candidate term word]
24. key behaviors change inventory.mp. [mp=title, abstract, heading word, drug trade name, original title, device manufacturer, drug manufacturer, device trade name, keyword heading word, floating subheading word, candidate
25. KBCI.mp. [mp=title, abstract, heading word, drug trade name, original title, device manufacturer, drug manufacturer, device trade name, keyword heading word, floating subheading word, candidate term word]
26. apathy evaluation scale.mp. [mp=title, abstract, heading word, drug trade name, original title, device manufacturer, drug manufacturer, device trade name, keyword heading word, floating subheading word, candidate term word]
27. apathy scale.mp. [mp=title, abstract, heading word, drug trade name, original title, device manufacturer, drug manufacturer, device trade name, keyword heading word, floating subheading word, candidate term word]
28. irritability apathy scale.mp. [mp=title, abstract, heading word, drug trade name, original title, device manufacturer, drug manufacturer, device trade name, keyword heading word, floating subheading word, candidate term word]
29. IAS.mp. [mp=title, abstract, heading word, drug trade name, original title, device manufacturer, drug manufacturer, device trade name, keyword heading word, floating subheading word, candidate term word]
30. 9 or 10 or 11 or 12 or 13 or 14 or 15 or 16 or 17 or 18 or 19 or 20 or 21 or 22 or 23 or 24 or 25 or 26 or 27 or 28 or 29 31. 8 and 30

**Supplemental Material 2.** Model underlying Rücker’s limit meta-analysis method.

The method by Rücker et al. (2011) computes an effect estimate adjusted for bias in meta-analysis. The underlying model is an extended random-effects model that takes account of possible small study effects by allowing the treatment effect to depend systematically on the standard error:

theta(i) = beta + sqrt(SE(i)^2^ + tau^2^)(epsilon(i) + alpha),

where theta(i) is the observed effect in study i, beta is the global mean, SE(i) the within-study standard error, tau^2^ the between-study variance, epsilon(i) follows a standard normal distribution, and alpha is the bias introduced by small-study effects.

**Supplemental R Script 1**. R script for the calculation of Pearson’s r estimate based on the degrees of freedom (sample size - 2) and exact p-value, when the direction of the relationship is known.

\# Required libraries library(MASS)

\# Target p-value target_p_value <-0.0009

\# Degrees of freedom (sample size - 2) df <- 2963

\# Calculate the critical value for t

t_critical <- qt(1 - target_p_value / 2, df = df)

\# Convert t to r

r_critical <- sqrt(t_critical^2 / (t_critical^2 + df))

\# Print the critical value for r print(r_critical)

\# Sample size n <- 45

\# Pearson correlation coefficient r <- -0.25

\# Compute the t-statistic

t_statistic <- r * sqrt((n - 2) / (1 - r^2))

\# Degrees of freedom df <- n - 2

\# Compute the p-value

p_value <- 2 * pt(abs(t_statistic), df = df, lower.tail = FALSE)

\# Print the p-value print(p_value)

---

**Supplemental Figure 1.**
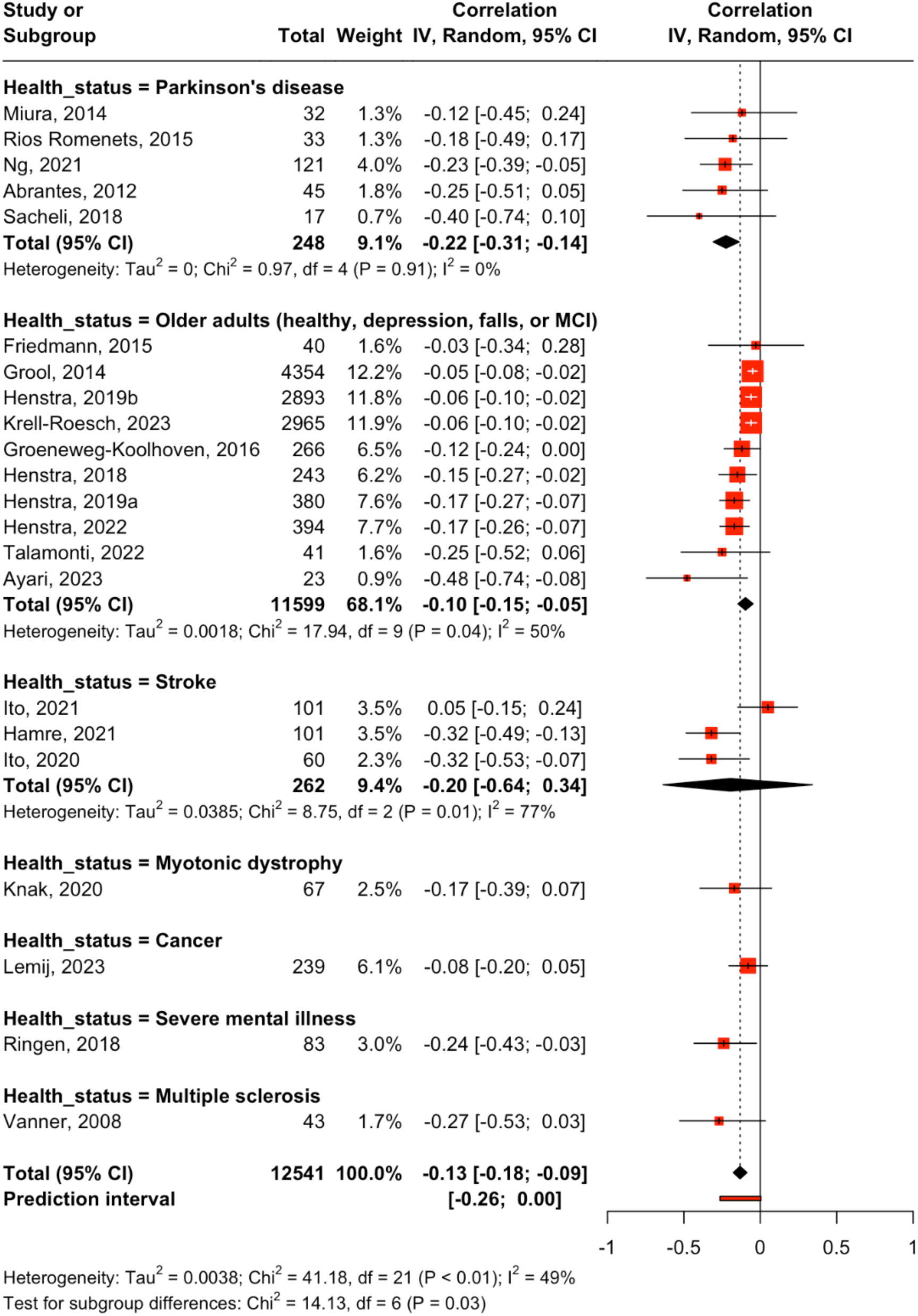
Subgroup meta-analyses. Differences according to health status. CI = confidence interval, IV = inverse variance, MCI = mild cognitive impairment.

**Supplemental Figure 2.**
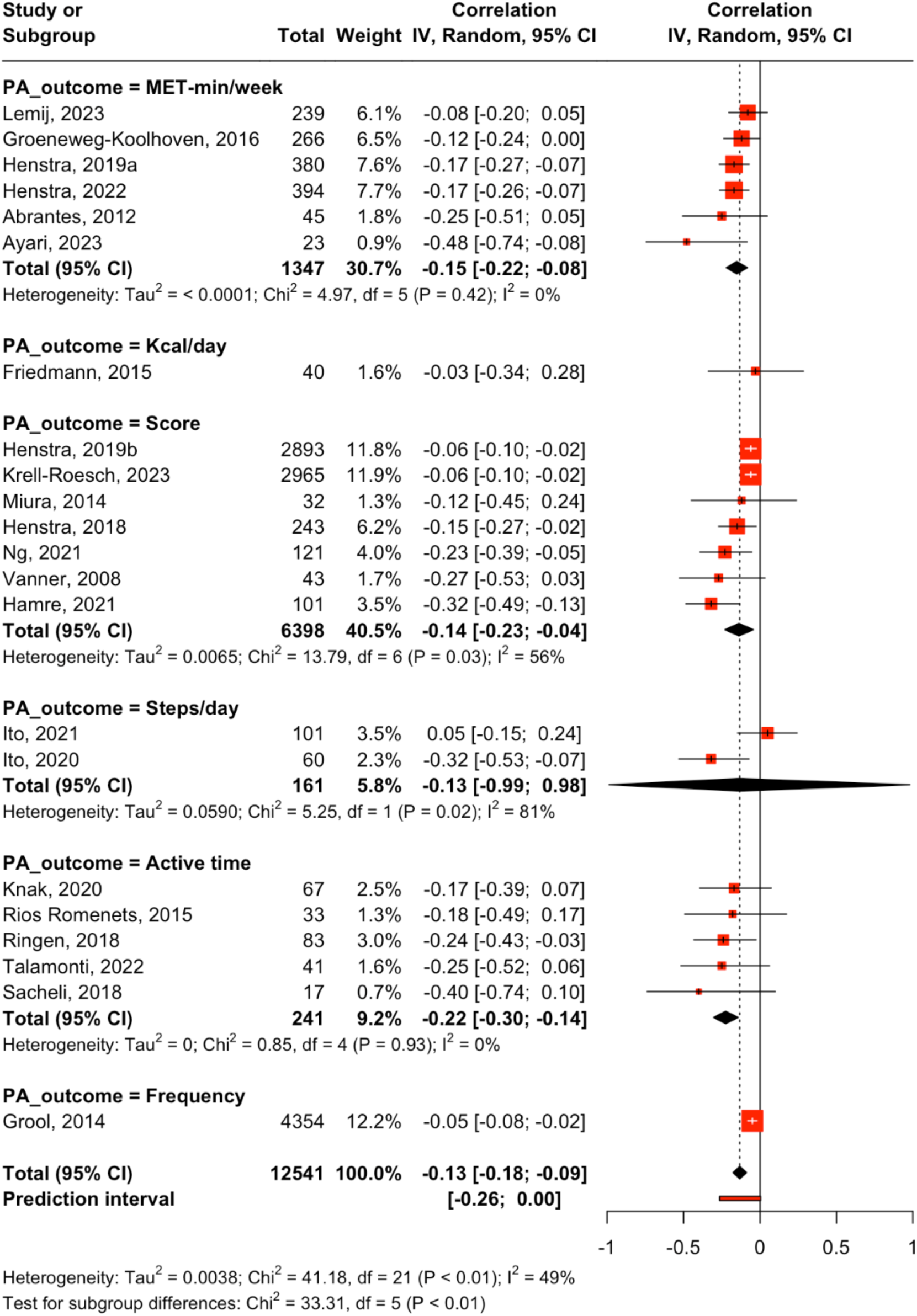
Subgroup meta-analyses. Differences according to physical activity outcome. CI = confidence interval, IV = inverse variance, PA = physical activity.

**Supplemental Figure 3.**
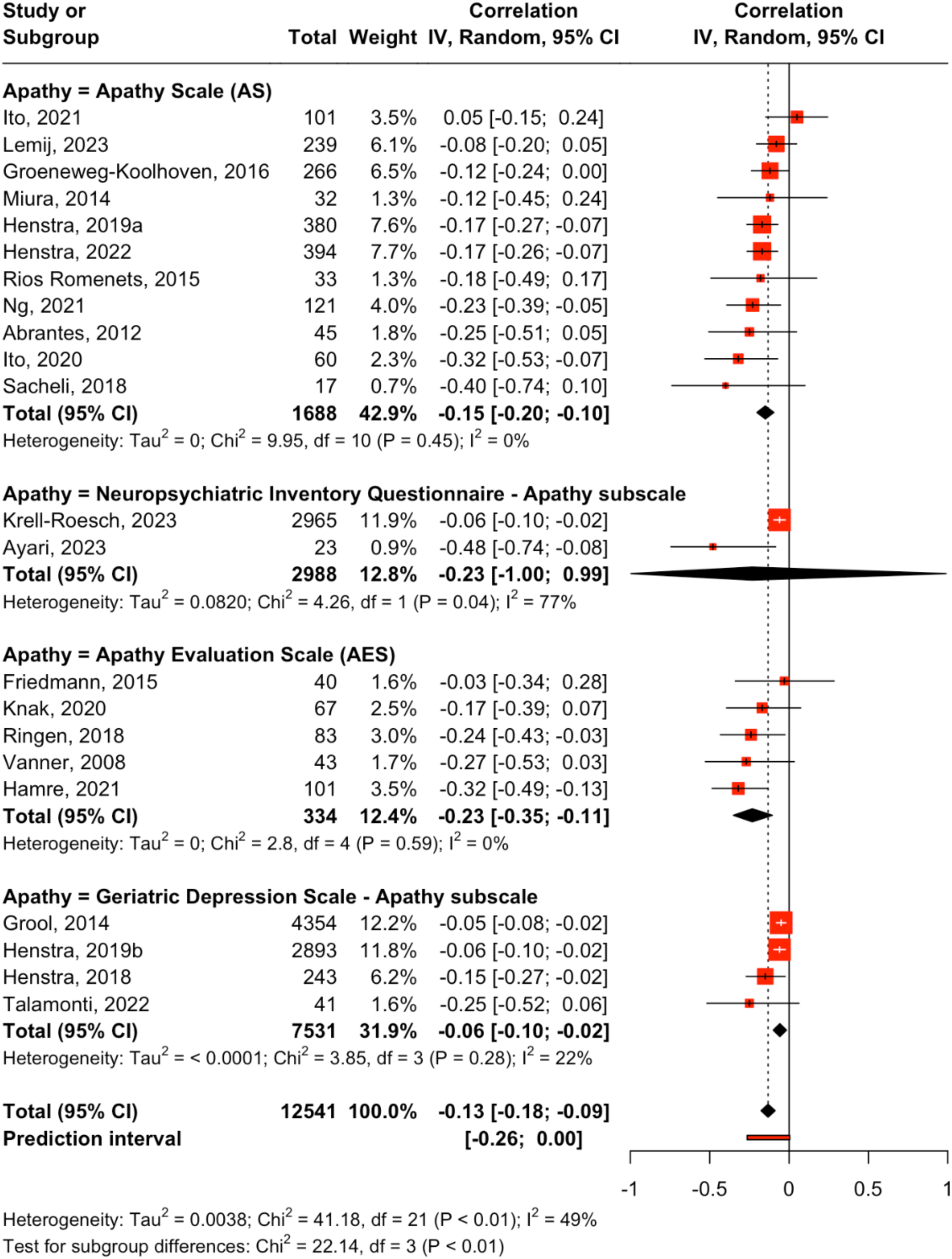
Subgroup meta-analyses. Differences according to apathy measure. CI = confidence interval, IV = inverse variance.

## References

1. Elliott TR, Warren AM. Why psychology is important in rehabilitation. In: Kennedy P, ed. Psychological management of physical disabilities: a practitioner’s guide. London: Routledge/Taylor & Francis Group, 2007:14–39.

2. Vallerand RJ, Thill EE. Introduction au concept de motivation. In: Vallerand RJ, Thill EE, eds. Introduction à la psychologie de la motivation. Laval, QC: Études Vivantes; 1993:3–40. [French]

3. Rhodes RE, Fiala B. Building motivation and sustainability into the prescription and recommendations for physical activity and exercise therapy: the evidence. Physiother Theory Pract. 2009;25(5-6):424–41. 10.1080/09593980902835344

4. Tay J, Morris RG, Markus HS. Apathy after stroke: diagnosis, mechanisms, consequences, and treatment. Int J Stroke. 2021;16(5):510–518. 10.1177/1747493021990906

5. Marin RS. Differential diagnosis and classification of apathy. Am J Psychiatry. 1990;147(1):22–30. 10.1176/ajp.147.1.22

6. Marin RS. Apathy: a neuropsychiatric syndrome. J Neuropsychiatry Clin Neurosci. 1991;3(3):243-254. 10.1176/jnp.3.3.243

7. Robert P, Onyike CU, Leentjens AF, et al. Proposed diagnostic criteria for apathy in Alzheimer’s disease and other neuropsychiatric disorders. Eur Psychiatry. 2009;24(2):98–104. 10.1016/j.eurpsy.2008.09.001

8. Costello H, Husain M, Roiser JP. Apathy and motivation: biological basis and drug treatment. Annu Rev Pharmacol Toxicol. 2024;64:313–338. 10.1146/annurev-pharmtox-022423-014645

9. Levy R, Dubois B. Apathy and the functional anatomy of the prefrontal cortex-basal ganglia circuits. Cereb Cortex. 2006;16(7):916–928. 10.1093/cercor/bhj043

10. Dickson SS, Husain M. Are there distinct dimensions of apathy? The argument for reappraisal. Cortex. 2022;149:246–256. 10.1016/j.cortex.2022.01.001

11. Le Heron C, Apps MAJ, Husain M. The anatomy of apathy: a neurocognitive framework for amotivated behaviour. Neuropsychologia. 2018;118(Pt B):54–67. 10.1016/j.neuropsychologia.2017.07.003

12. Leung DKY, Chan WC, Spector A, Wong GHY. Prevalence of depression, anxiety, and apathy symptoms across dementia stages: a systematic review and meta-analysis. Int J Geriatr Psychiatry. 2021;36(9):1330–1344. 10.1002/gps.5556

13. Mulin E, Leone E, Dujardin K, et al. Diagnostic criteria for apathy in clinical practice. Int J Geriatr Psychiatry. 2011;26(2):158–165. 10.1002/gps.2508

14. Yazbek H, Norton J, Capdevielle D, et al. The Lille Apathy Rating Scale (LARS): exploring its psychometric properties in schizophrenia. Schizophr Res. 2014;157(1-3):278–284. 10.1016/j.schres.2014.04.034

15. Faerden A, Finset A, Friis S, et al. Apathy in first episode psychosis patients: one year follow up. Schizophr Res. 2010;116(1):20–26. 10.1016/j.schres.2009.10.014

16. den Brok MG, van Dalen JW, van Gool WA, Moll van Charante EP, de Bie RM, Richard E. Apathy in Parkinson’s disease: a systematic review and meta-analysis. Mov Disord. 2015;30(6):759–769. 10.1002/mds.26208

17. Zhang H, Feng Y, Lv H, Tang S, Peng Y. The prevalence of apathy in stroke patients: a systematic review and meta-analysis. J Psychosom Res. 2023;173:111478. 10.1016/j.jpsychores.2023.111478

18. Pardini M, Cordano C, Guida S, et al. Prevalence and cognitive underpinnings of isolated apathy in young healthy subjects. J Affect Disord. 2016;189:272–275. 10.1016/j.jad.2015.09.062

19. Hwang TJ, Masterman DL, Ortiz F, Fairbanks LA, Cummings JL. Mild cognitive impairment is associated with characteristic neuropsychiatric symptoms. Alzheimer Dis Assoc Disord. 2004;18(1):17–21. 10.1097/00002093-200401000-00004

20. Onyike CU, Sheppard JM, Tschanz JT, et al. Epidemiology of apathy in older adults: the Cache County Study. Am J Geriatr Psychiatry. 2007;15(5):365–375. 10.1097/01.JGP.0000235689.42910.0d

21. Geda YE, Roberts RO, Knopman DS, et al. Prevalence of neuropsychiatric symptoms in mild cognitive impairment and normal cognitive aging: population-based study. Arch Gen Psychiatry. 2008;65(10):1193–1198. 10.1001/archpsyc.65.10.1193

22. Lyketsos CG, Steinberg M, Tschanz JT, Norton MC, Steffens DC, Breitner JC. Mental and behavioral disturbances in dementia: findings from the Cache County Study on Memory in Aging. Am J Psychiatry. 2000;157(5):708–714. 10.1176/appi.ajp.157.5.708

23. Groeneweg-Koolhoven I, de Waal MW, van der Weele GM, Gussekloo J, van der Mast RC. Quality of life in community-dwelling older persons with apathy. Am J Geriatr Psychiatry. 2014;22(2):186–194. 10.1016/j.jagp.2012.10.024

24. Pink A, Stokin GB, Bartley MM, et al. Neuropsychiatric symptoms, APOE ε4, and the risk of incident dementia: a population-based study. Neurology. 2015;84(9):935–943. 10.1212/WNL.0000000000001307

25. Guercio BJ, Donovan NJ, Munro CE, et al. The Apathy Evaluation Scale: a comparison of subject, informant, and clinician report in cognitively normal elderly and mild cognitive impairment. J Alzheimers Dis. 2015;47(2):421–432. 10.3233/JAD-150146

26. Mega MS, Cummings JL, Fiorello T, Gornbein J. The spectrum of behavioral changes in Alzheimer’s disease. Neurology. 1996;46(1):130–135. 10.1212/wnl.46.1.130

27. Lanctôt KL, Agüera-Ortiz L, Brodaty H, et al. Apathy associated with neurocognitive disorders: recent progress and future directions. Alzheimers Dement. 2017;13(1):84–100. 10.1016/j.jalz.2016.05.008

28. Ayers E, Shapiro M, Holtzer R, Barzilai N, Milman S, Verghese J. Symptoms of apathy independently predict incident frailty and disability in community-dwelling older adults. J Clin Psychiatry. 2017;78(5):e529–e536. 10.4088/JCP.15m10113

29. Clarke DE, Ko JY, Lyketsos C, Rebok GW, Eaton WW. Apathy and cognitive and functional decline in community-dwelling older adults: results from the Baltimore ECA longitudinal study. Int Psychogeriatr. 2010;22(5):819–829. 10.1017/S1041610209991402

30. Wadsworth LP, Lorius N, Donovan NJ, et al. Neuropsychiatric symptoms and global functional impairment along the Alzheimer’s continuum. Dement Geriatr Cogn Disord. 2012;34(2):96–111. 10.1159/000342119

31. Musa Salech G, Lillo P, van der Hiele K, Méndez-Orellana C, Ibáñez A, Slachevsky A. Apathy, executive function, and emotion recognition are the main drivers of functional impairment in behavioral variant of frontotemporal dementia. Front Neurol. 2022;12:734251. 10.3389/fneur.2021.734251

32. Ishii S, Weintraub N, Mervis JR. Apathy: a common psychiatric syndrome in the elderly. J Am Med Dir Assoc. 2009;10(6):381–393. 10.1016/j.jamda.2009.03.007

33. Barone P, Antonini A, Colosimo C, et al. The PRIAMO study: a multicenter assessment of nonmotor symptoms and their impact on quality of life in Parkinson’s disease. Mov Disord. 2009;24(11):1641–1649. 10.1002/mds.22643

34. Vilalta-Franch J, Calvó-Perxas L, Garre-Olmo J, Turró-Garriga O, López-Pousa S. Apathy syndrome in Alzheimer’s disease epidemiology: prevalence, incidence, persistence, and risk and mortality factors. J Alzheimers Dis. 2013;33(2):535–543. 10.3233/JAD-2012-120913

35. Lansdall CJ, Coyle-Gilchrist ITS, Vázquez Rodríguez P, et al. Prognostic importance of apathy in syndromes associated with frontotemporal lobar degeneration. Neurology. 2019;92(14):e1547–e1557. 10.1212/WNL.0000000000007249

36. Kruse C, Maier F, Spottke A, et al. Apathy in patients with Alzheimer’s disease is a cost-driving factor. Alzheimers Dement. 2023;19(7):2853–2864. 10.1002/alz.12915

37. Vanner EA, Block P, Christodoulou CC, Horowitz BP, Krupp LB. Pilot study exploring quality of life and barriers to leisure-time physical activity in persons with moderate to severe multiple sclerosis. Disabil Health J. 2008;1(1):58–65. 10.1016/j.dhjo.2007.11.001

38. Yao H, Takashima Y, Araki Y, Uchino A, Yuzuriha T, Hashimoto M. Leisure-time physical inactivity associated with cascular depression or apathy in community-dwelling elderly subjects: the Sefuri Study. J Stroke Cerebrovasc Dis. 2015;24(11):2625–2631. 10.1016/j.jstrokecerebrovasdis.2015.07.018

39. Ishimaru D, Tanaka H, Nagata Y, Takabatake S, Nishikawa T. Physical activity in severe dementia is associated with agitation rather than cognitive function. Am J Alzheimers Dis Other Demen. 2020;35:1533317519871397. 10.1177/1533317519871397

40. Ng SY, Chia NS, Abbas MM, et al. Physical activity improves anxiety and apathy in early Parkinson’s disease: a longitudinal follow-up study. Front Neurol. 2021;11:625897. 10.3389/fneur.2020.625897

41. Bull FC, Al-Ansari SS, Biddle S, et al. World Health Organization 2020 guidelines on physical activity and sedentary behaviour. Br J Sports Med. 2020;54(24):1451–1462. 10.1136/bjsports-2020-102955

42. Cheval B, Darrous L, Choi KW, et al. Genetic insights into the causal relationship between physical activity and cognitive functioning. Sci Rep. 2023;13(1):5310. 10.1038/s41598-023-32150-1

43. Wahid A, Manek N, Nichols M, et al. Quantifying the association between physical activity and cardiovascular disease and diabetes: a systematic review and meta-analysis. J Am Heart Assoc. 2016;5(9):e002495. 10.1161/JAHA.115.002495

44. Moore SC, Lee IM, Weiderpass E, et al. Association of leisure-time physical activity with risk of 26 types of cancer in 1.44 million adults. JAMA Intern Med. 2016;176(6):816–825. 10.1001/jamainternmed.2016.1548

45. Liu X, Zhang D, Liu Y, et al. Dose-response association between physical activity and incident hypertension: a systematic review and meta-analysis of cohort studies. Hypertension. 2017;69(5):813–820. 10.1161/HYPERTENSIONAHA.116.08994

46. Cheval B, Maltagliati S, Sieber S, et al. Why are individuals with diabetes less active? The mediating role of physical, emotional, and cognitive factors. Ann Behav Med. 2021;55(9):904–917. 10.1093/abm/kaaa120

47. Bleich SN, Vercammen KA, Zatz LY, Frelier JM, Ebbeling CB, Peeters A. Interventions to prevent global childhood overweight and obesity: a systematic review. Lancet Diabetes Endocrinol. 2018;6(4):332–346. 10.1016/S2213-8587(17)30358-3

48. Boisgontier MP, Orsholits D, von Arx M, et al. Adverse childhood experiences, depressive symptoms, functional dependence, and physical activity: a moderated mediation model. J Phys Act Health. 2020;17(8):790–799. 10.1123/jpah.2019-0133

49. van Allen ZM, Orsholits D, Boisgontier MP. Prestroke physical activity matters for functional limitations: a longitudinal case-control study of 12,860 participants. Phys Ther. 2024;104(10):pzae094. 10.1093/ptj/pzae094

50. van Allen ZM, Boisgontier MP. Prospective classification of functional dependence: insights from machine learning and the Canadian Longitudinal Study on Aging. MedRxiv. 10.1101/2024.07.15.24310429

51. Knak KL, Sheikh AM, Witting N, Vissing J. Physical activity in myotonic dystrophy type 1. J Neurol. 2020;267(6):1679–1686. 10.1007/s00415-020-09758-8

52. Ito D, Kawakami M, Narita Y, Yoshida T, Mori N, Kondo K. Cognitive function is a predictor of the daily step count in patients with subacute stroke with independent walking ability: a prospective cohort study. Arch Rehabil Res Clin Transl. 2021;3(3):100132. 10.1016/j.arrct.2021.100132

53. Ersöz Hüseyinsinoğlu B, Kuran Aslan G, Tarakci D, Razak Özdinçler A, Küçükoğlu H, Baybaş S. Physical activity level of ambulatory stroke patients: is it related to neuropsychological factors? Noro Psikiyatr Ars. 2017;54(2):155–161. 10.5152/npa.2016.12760

54. Cheval B, Sieber S, Guessous I, et al. Effect of early-and adult-life socioeconomic circumstances on physical inactivity. Med Sci Sports Exerc. 2018;50(3):476–485. 10.1249/MSS.0000000000001472

55. Brodaty H, Altendorf A, Withall A, Sachdev P. Do people become more apathetic as they grow older? A longitudinal study in healthy individuals. Int Psychogeriatr. 2010;22(3):426–436. 10.1017/S1041610209991335

56. Spalletta G, Fagioli S, Caltagirone C, Piras F. Brain microstructure of subclinical apathy phenomenology in healthy individuals. Hum Brain Mapp. 2013;34(12):3193–3203. 10.1002/hbm.22137

57. Page MJ, Moher D, Bossuyt PM, et al. PRISMA 2020 explanation and elaboration: updated guidance and exemplars for reporting systematic reviews. BMJ. 2021;372:n160. 10.1136/bmj.n160

58. Farajzadeh A, Hebert A, Bilodeau M, Boisgontier M. Apathy and physical activity: a systematic review. PROSPERO (CRD42023492162) [preregistration]; December 21, 2023. Available from: https://www.crd.york.ac.uk/prospero/display_record.php?ID=CRD42023492162

59. Goubran M, Farajzadeh A, Lahart IM, Bilodeau M, Boisgontier MP. Relationship between fear of movement and physical activity in patients with cardiac, rheumatologic, neurologic, pulmonary, or pain conditions: A systematic review and meta-analysis. Phys Ther. (In press)

60. Hutchinson J. Evidence of the association between kinesiophobia and physical inactivity. Peer Community In Health Mov Sci. 2024:100039. 10.24072/pci.healthmovsci.100039

61. National Institutes of Health. Quality assessment tool for observational cohort and cross-sectional studies [Internet]. 2014 (accessed online September 2024). https://www.nhlbi.nih.gov/health-topics/study-quality-assessment-tools

62. R Core Team. R: A language and environment for statistical computing [Computer software]. Version 4.3.1. Vienna, Austria: Foundation for Statistical Computing; 2023. https://www.r-project.org

63. Schwarzer G. meta: general package for meta-analysis [R package]. Version 6.5-0; 2023. https://cran.rproject.org/web/packages/meta/meta.pdf

64. Schwarzer G, Carpenter RJ, Rücker G. *metasens: advanced statistical methods to model and adjust for bias in meta-analysis* [R package]. Version 1.5-2; 2023. https://cran.rproject.org/web/packages/metasens/metasens.pdf

65. Viechtbauer W. Conducting meta-analyses in R with the metafor package. J Stat Softw. 2010;36(3):1–48. https://cran.r-project.org/web/packages/metafor/metafor.pdf

66. Viechtbauer W. metafor: meta-analysis package for R [R package]. Version 4.2-0; 2023. https://cran.r-project.org/web/packages/metafor/metafor.pdf

67. Ripley B, Venables B, Bates DM, Hornik K, Gebhardt A, Firth D. *MASS: Support Functions and Datasets for Venables and Ripley’s MASS* [R package]. Version 7.3–61; 2024. https://cran.r-project.org/web/packages/MASS/MASS.pdf

68. Cohen J, Cohen P, West SG, Aiken LS. Applied multiple regression/correlation analysis for the behavioral sciences. 3rd ed. Mahwah, NJ: Lawrence Erlbaum Associates; 2002.

69. Viechtbauer W. Bias and efficiency of metanalytic variance estimators in the random effects model. J Educ Behav Stat. 2005;30(3):261–293. 10.3102/1076998603000326

70. Higgins JPT, Thompson SG. Quantifying heterogeneity in a meta-analysis. Stat Med. 2002;21(11):1539–1558. 10.1002/sim.1186

71. Knapp G, Hartung J. Improved tests for a random effects meta-regression with a single covariate. Stat Med. 2003;22(17):2693–2710. 10.1002/sim.1482

72. Cohen J. Statistical power analysis for the behavioral sciences. 2nd ed.; 1988.

73. Lin L, Chu H. Quantifying publication bias in meta-analysis. Biometrics. 2018;74(3):785–794. 10.1111/biom.12817

74. Egger M, Davey Smith G, Schneider M, Minder C. Bias in meta-analysis detected by a simple, graphical test. BMJ. 1997;315(7109):629-634. 10.1136/bmj.315.7109.629

75. Rücker G, Schwarzer G, Carpenter JR, Binder H, Schumacher M. Treatment-effect estimates adjusted for small-study effects via a limit meta-analysis. Biostatistics. 2011;12(1):122–142. 10.1093/biostatistics/kxq046

76. Simonsohn U, Nelson LD, Simmons JP. P-curve: a key to the file-drawer. J Exp Psychol Gen. 2014;143(2):534–547. 10.1037/a0033242

77. Cheung M. Modeling dependent effect sizes with three-level meta-analyses: a structural equation modeling approach. Psychol Methods. 2014;19(2):211–229. 10.1037/a0032968

78. Ayari S, Abellard A, Sakrani S, et al. Comparison of dance and aerobic exercise on cognition and neuropsychiatric symptoms in sedentary older adults with cognitive impairment. Eur Geriatr Med. 2023;14(6):1289–1299. 10.1007/s41999-023-00849-z

79. Friedmann E, Galik E, Thomas SA, Hall PS, Chung SY, McCune S. Evaluation of a pet-assisted living intervention for improving functional status in assisted living residents with mild to moderate cognitive impairment: a pilot study. Am J Alzheimers Dis Other Demen. 2015;30(3):276–289. 10.1177/1533317514545477

80. Di Santo SG, Franchini F, Filiputti B, Martone A, Sannino S. The effects of COVID-19 and quarantine measures on the lifestyles and mental health of people over 60 at increased risk of dementia. Front Psychiatry. 2020;11:578628. 10.3389/fpsyt.2020.578628

81. Trumpf R, Haussermann P, Zijlstra W, Fleiner T. Circadian aspects of mobility-related behavior in patients with dementia: an exploratory analysis in acute geriatric psychiatry. Int J Geriatr Psychiatry. 2023;38(6):e5957. 10.1002/gps.5957

82. Galik E, Resnick B, Hammersla M, Brightwater J. Optimizing function and physical activity among nursing home residents with dementia: testing the impact of function-focused care. Gerontologist. 2014;54(6):930–943. 10.1093/geront/gnt108

83. Arbour-Nicitopoulos KP, Duncan MJ, Remington G, Cairney J, Faulkner GE. The utility of the health action process approach model for predicting physical activity intentions and behavior in schizophrenia. Front Psychiatry. 2017;8:135. 10.3389/fpsyt.2017.00135

84. Rios Romenets S, Anang J, Fereshtehnejad SM, Pelletier A, Postuma R. Tango for treatment of motor and non-motor manifestations in Parkinson’s disease: a randomized control study. Complement Ther Med. 2015;23(2):175–184. 10.1016/j.ctim.2015.01.015

85. Groeneweg-Koolhoven I, Comijs HC, Naarding P, de Waal MW, van der Mast RC. Apathy in older persons with depression: course and predictors: the NESDO Study. J Geriatr Psychiatry Neurol. 2016;29(4):178–186. 10.1177/0891988716632914

86. Gorzkowska A, Cholewa J, Małecki A, Klimkowicz-Mrowiec A, Cholewa J. What determines spontaneous physical activity in patients with Parkinson’s disease? J Clin Med. 2020;9(5):1296. 10.3390/jcm9051296

87. Ito D, Tanaka T, Kunieda Y, et al. Factors associated with post-stroke apathy in subacute stroke patients. Psychogeriatrics. 2020;20(5):780–781. 10.1111/psyg.12551

88. Hamre C, Fure B, Helbostad JL, et al. Factors associated with level of physical activity after minor stroke. J Stroke Cerebrovasc Dis. 2021;30(4):105628. 10.1016/j.jstrokecerebrovasdis.2021.105628

89. Abrantes AM, Friedman JH, Brown RA, et al. Physical activity and neuropsychiatric symptoms of Parkinson disease. J Geriatr Psychiatry Neurol. 2012;25(3):138–145. 10.1177/0891988712455237

90. Sacheli MA, Murray DK, Vafai N, et al. Habitual exercisers versus sedentary subjects with Parkinson’s disease: multimodal PET and fMRI study. Mov Disord. 2018;33(12):1945–1950. 10.1002/mds.27498

91. Miura K, Takashima S, Matsui M, Tanaka K. Low frequency of leisure-time activities correlates with cognitive decline and apathy in patients with Parkinson’s disease. Adv Parkinsons Dis. 2014;3:15–21. 10.4236/apd.2014.33004

92. Farholm A, Sørensen M, Halvari H, Hynnekleiv T. Associations between physical activity and motivation, competence, functioning, and apathy in inhabitants with mental illness from a rural municipality: a cross-sectional study. BMC Psychiatry. 2017;17(1):359. 10.1186/s12888-017-1528-3

93. Ringen PA, Faerden A, Antonsen B, et al. Cardiometabolic risk factors, physical activity and psychiatric status in patients in long-term psychiatric inpatient departments. Nord J Psychiatry. 2018;72(4):296–302. 10.1080/08039488.2018.1449012

94. Lemij AA, Liefers GJ, Derks MGM, et al. Physical function and physical activity in older breast cancer survivors: 5-year follow-up from the Climb Every Mountain Study. Oncologist. 2023;28(6):e317-e323. 10.1093/oncolo/oyad027

95. Henstra MJ, Feenstra TC, van der Velde N, et al. Apathy is associated with greater decline in subjective, but not in objective measures of physical functioning in older people without dementia. J Gerontol A Biol Sci Med Sci. 2019a;74(2):254–260. 10.1093/gerona/gly014

96. Henstra M, Giltay E, van der Mast R, van der Velde N, Rhebergen D, Rius Ottenheim N. Does late-life depression counteract the beneficial effect of physical activity on cognitive decline? Results from the NESDO Study. J Geriatr Psychiatry Neurol. 2022;35(3):450–459. 10.1177/08919887211002658

97. Krell-Roesch J, Syrjanen JA, Bezold J, et al. Mid- and late-life physical activity and neuropsychiatric symptoms in dementia-free older adults: Mayo Clinic Study of Aging. J Neuropsychiatry Clin Neurosci. 2023;35(2):133–140. 10.1176/appi.neuropsych.20220068

98. Henstra MJ, Houbolt CM, Seppala LJ, et al. Age modifies the association between apathy and recurrent falling in Dutch ambulant older persons with a high fall risk: recurrent falling in Dutch outpatients, does apathy play a role? Exp Gerontol. 2018;112:54–62. 10.1016/j.exger.2018.09.002

99. Henstra MJ, Rhebergen D, Stek ML, et al. The association between apathy, decline in physical performance, and falls in older persons. Aging Clin Exp Res. 2019b;31(10):1491–1499. 10.1007/s40520-018-1096-5

100. Hashimoto M, Araki Y, Takashima Y, et al. Hippocampal atrophy and memory dysfunction associated with physical inactivity in community-dwelling elderly subjects: the Sefuri Study. Brain Behav. 2016;7(2):e00620. 10.1002/brb3.620

101. Grool AM, Geerlings MI, Sigurdsson S, et al. Structural MRI correlates of apathy symptoms in older persons without dementia: AGES-Reykjavik Study. Neurology. 2014;82(18):1628–1635. 10.1212/WNL.0000000000000378

102. Talamonti D, Dupuy EG, Boudaa S, et al. Prefrontal hyperactivation during dual-task walking related to apathy symptoms in older individuals. PLoS One. 2022;17(4):e0266553. 10.1371/journal.pone.0266553

103. Starkstein SE, Mayberg HS, Preziosi TJ, Andrezejewski P, Leiguarda R, Robinson RG. Reliability, validity, and clinical correlates of apathy in Parkinson’s disease. J Neuropsychiatry Clin Neurosci. 1992;4(2):134–139. 10.1176/jnp.4.2.134

104. Marin RS, Biedrzycki RC, Firinciogullari S. Reliability and validity of the Apathy Evaluation Scale. Psychiatry Res. 1991;38(2):143–162. 10.1016/0165-1781(91)90040-v

105. Craig CL, Marshall AL, Sjöström M, et al. International physical activity questionnaire: 12-country reliability and validity. Med Sci Sports Exerc. 2003;35(8):1381–1395. 10.1249/01.MSS.0000078924.61453.FB

106. Simonsohn U, Simmons JP, Nelson LD. Better P-curves: making P-curve analysis more robust to errors, fraud, and ambitious P-hacking, a reply to Ulrich and Miller (2015). J Exp Psychol Gen. 2015;144(6):1146–1152. 10.1037/xge0000104

107. Taylor RS, Walker S, Smart NA, et al. Impact of exercise rehabilitation on exercise capacity and quality-of-life in heart failure: individual participant meta-analysis. J Am Coll Cardiol. 2019;73(12):1430–1443. doi:10.1016/j.jacc.2018.12.072

108. Molloy C, Long L, Mordi IR, et al. Exercise-based cardiac rehabilitation for adults with heart failure. Cochrane Database Syst Rev. 2024;3(3):CD003331. doi:10.1002/14651858.CD003331.pub6

109. Turan TN, Nizam A, Lynn MJ, et al. Relationship between risk factor control and vascular events in the SAMMPRIS trial. Neurology. 2017;88(4):379–385. doi:10.1212/WNL.0000000000003534

110. Schuch F, Vancampfort D, Firth J, et al. Physical activity and sedentary behavior in people with major depressive disorder: a systematic review and meta-analysis. J Affect Disord. 2017;210:139–150. 10.1016/j.jad.2016.10.050

111. Boisgontier MP, Orsholits D, von Arx M, et al. Adverse childhood experiences, depressive symptoms, functional dependence, and physical activity: a moderated mediation model. J Phys Act Health. 2020;17(8):790–799. 10.1123/jpah.2019-0133

112. Bleich SN, Vercammen KA, Zatz LY. Interventions to prevent global childhood overweight and obesity: a systematic review. Lancet Diabetes Endocrinol. 2018;6:332–346. 10.1016/S2213-8587(17)30358-3

113. Najar J, Östling S, Gudmundsson P, et al. Cognitive and physical activity and dementia: a 44-year longitudinal population study of women. Neurology. 2019;92(12):E1322–E1330. 10.1212/WNL.0000000000007021

114. Daste C, Kirren Q, Akoum J, Lefèvre-Colau MM, Rannou F, Nguyen C. Physical activity for osteoarthritis: efficiency and review of recommandations. Joint Bone Spine. 2021;88(6):105207. 10.1016/j.jbspin.2021.105207

115. Brenner H, Chen C. The colorectal cancer epidemic: challenges and opportunities for primary, secondary and tertiary prevention. Br J Cancer. 2018;119(7):785–792. 10.1038/s41416-018-0264-x

116. Cai Y, Li L, Xu C, Wang Z. The effectiveness of non-pharmacological interventions on apathy in patients with dementia: a systematic review of systematic reviews. Worldviews Evid Based Nurs. 2020;17(4):311–318. 10.1111/wvn.12459

117. Cugusi L, Solla P, Serpe R, et al. Effects of a Nordic Walking program on motor and non-motor symptoms, functional performance and body composition in patients with Parkinson’s disease. NeuroRehabilitation. 2015;37(2):245–254. 10.3233/NRE-151257

118. King LA, Wilhelm J, Chen Y, et al. Effects of group, individual, and home exercise in persons with Parkinson disease: a randomized clinical trial. J Neurol Phys Ther. 2015;39(4):204–212. 10.1097/NPT.0000000000000101

119. Farajzadeh A, Jabouille F, Benoit N, Bezeau O, Bourgie T, Gerro B, Ouimet J, Boisgontier MP. Apathy, intentions, explicit attitudes, and approach-avoidance tendencies in physical activity behavior. MedRxiv. 10.1101/2024.07.16.24310493

120. Pessiglione M, Vinckier F, Bouret S, Daunizeau J, Le Bouc R. Why not try harder? Computational approach to motivation deficits in neuro-psychiatric diseases. Brain. 2018;141(3):629–650. 10.1093/brain/awx278

121. Remy P, Doder M, Lees A, Turjanski N, Brooks D. Depression in Parkinson’s disease: loss of dopamine and noradrenaline innervation in the limbic system. Brain. 2005;128(Pt 6):1314–1322. 10.1093/brain/awh445

122. Thobois S, Ardouin C, Lhommée E, et al. Non-motor dopamine withdrawal syndrome after surgery for Parkinson’s disease: predictors and underlying mesolimbic denervation. Brain. 2010;133(Pt 4):1111–1127. 10.1093/brain/awq032

123. Brown CA, Campbell MC, Karimi M, et al. Dopamine pathway loss in nucleus accumbens and ventral tegmental area predicts apathetic behavior in MPTP-lesioned monkeys. Exp Neurol. 2012;236(1):190–197. 10.1016/j.expneurol.2012.04.025

124. Martínez-Horta S, Riba J, de Bobadilla RF, et al. Apathy in Parkinson’s disease: neurophysiological evidence of impaired incentive processing. J Neurosci. 2014;34(17):5918–5926. 10.1523/JNEUROSCI.0251-14.2014

125. Muhammed K, Manohar S, Ben Yehuda M, et al. Reward sensitivity deficits modulated by dopamine are associated with apathy in Parkinson’s disease. Brain. 2016;139(Pt 10):2706–2721. 10.1093/brain/aww188

126. Chong TT, Bonnelle V, Husain M. Quantifying motivation with effort-based decision-making paradigms in health and disease. Prog Brain Res. 2016;229:71–100. 10.1016/bs.pbr.2016.05.002

127. Porat O, Hassin-Baer S, Cohen OS, Markus A, Tomer R. Asymmetric dopamine loss differentially affects effort to maximize gain or minimize loss. Cortex. 2014;51:82–91. 10.1016/j.cortex.2013.10.004

128. Le Bouc R, Rigoux L, Schmidt L, et al. Computational dissection of dopamine motor and motivational functions in humans. J Neurosci. 2016;36(25):6623–6633. 10.1523/JNEUROSCI.3078-15.2016

129. Hortobágyi T, Finch A, Solnik S, Rider P, DeVita P. Association between muscle activation and metabolic cost of walking in young and old adults. J Gerontol A Biol Sci Med Sci. 2011;66(5):541–547. 10.1093/gerona/glr008

130. Martin PE, Rothstein DE, Larish DD. Effects of age and physical activity status on the speed-aerobic demand relationship of walking. J Appl Physiol. 1992;73(1):200–206. 10.1152/jappl.1992.73.1.200

131. Ortega JD, Farley CT. Effects of aging on mechanical efficiency and muscle activation during level and uphill walking. J Electromyogr Kinesiol. 2015;25(1):193–198. 10.1016/j.jelekin.2014.09.003

132. Harrison F, Mortby ME, Mather KA, Sachdev PS, Brodaty H. Apathy as a determinant of health behaviors in older adults: implications for dementia risk reduction. Alzheimers Dement. 2023;15(4):e12505. 10.1002/dad2.12505

133. Gabelica M, Bojčić R, Puljak L. Many researchers were not compliant with their published data sharing statement: a mixed-methods study. J Clin Epidemiol. 2022;150:33–41. 10.1016/j.jclinepi.2022.05.019

134. Boisgontier MP, Iversen MD. Physical inactivity: a behavioral disorder in the physical therapist’s scope of practice. Phys Ther. 2020;100(5):743–746. 10.1093/ptj/pzaa011

135. Boisgontier MP. Research integrity requires to be aware of good and questionable research practices. Eur Rehabil J. 2021;2(1):1–3. 10.52057/erj.v2i1.24

136. Lahart IM, Boisgontier MP. (2024). *Apathy and physical activity: data and Rmd script for meta-analysis*. [Data set, Rmd scripts, supplementary material]. Version 1.0; 2024. 10.5281/zenodo.10929857

137. Allen L, O’Connell A, Kiermer V. How can we ensure visibility and diversity in research contributions? How the contributor role taxonomy (CRediT) is helping the shift from authorship to contributorship. Learn Publ. 2019;32(1):71–74. 10.1002/leap.1210

## Reference

139. - Rücker G, Schwarzer G, Carpenter JR, Binder H, Schumacher M. Treatment-effect estimates adjusted for small-study effects via a limit meta-analysis. Biostatistics. 2011;12(1):122–142. 10.1093/biostatistics/kxq046

## References

141. - Ripley B, Venables B, Bates DM, Hornik K, Gebhardt A, Firth D. MASS: Support Functions and Datasets for Venables and Ripley’s MASS [R package]. Version 7.3-61; 2024. https://cran.r-project.org/web/packages/MASS/MASS.pdf

142. - Cohen J, Cohen P, West SG, Aiken LS. Applied multiple regression/correlation analysis for the behavioral sciences. 3rd ed. Mahwah, NJ: Lawrence Erlbaum Associates; 2002.”

## Reference

144. - Cohen J, Cohen P, West SG, Aiken LS. Applied multiple regression/correlation analysis for the behavioral sciences. 3rd ed. Mahwah, NJ: Lawrence Erlbaum Associates; 2002.”

